# Maternal Fc-mediated non-neutralizing antibody responses correlate with protection against congenital human cytomegalovirus infection

**DOI:** 10.1101/2021.12.05.21267312

**Authors:** Eleanor C. Semmes, Itzayana G. Miller, Jennifer A. Jenks, Courtney E. Wimberly, Stella J. Berendam, Melissa J. Harnois, Helen Webster, Jillian H. Hurst, Joanne Kurtzberg, Genevieve G Fouda, Kyle M. Walsh, Sallie R. Permar

**Affiliations:** Medical Scientist Training Program, Department of Molecular Genetics and Microbiology, Duke University, Durham, NC, USA; Duke Human Vaccine Institute, Duke University, Durham, NC, USA; Duke Children’s Health & Discovery Initiative, Duke University, Durham, NC, USA; Department of Neurosurgery, Duke University, Durham, NC, USA; Department of Pediatrics, Duke University, Durham, NC, USA; Carolinas Cord Blood Bank, Duke University Medical Center, Durham, NC, USA; Department of Pediatrics, Weill Cornell School of Medicine, New York City, NY, USA

## Abstract

Human cytomegalovirus (HCMV) is the most common congenital infection and a leading cause of stillbirth, neurodevelopmental impairment, and pediatric hearing loss worldwide. Development of a maternal vaccine or therapeutic to prevent congenital infection has been hindered by limited knowledge of the immune responses that protect against placental HCMV transmission in maternal primary and nonprimary infection. To identify protective antibody responses, we measured anti-HCMV IgG binding and anti-viral functions in maternal and cord blood sera from HCMV transmitting (n=41) and non-transmitting (n=40) mother-infant dyads identified via a large U.S.-based public cord blood bank. In a predefined immune correlate analysis, maternal monocyte-mediated antibody-dependent cellular phagocytosis (ADCP) and high avidity IgG binding to HCMV envelope glycoproteins were associated with decreased risk of congenital HCMV infection. Moreover, HCMV-specific IgG engagement of FcγRI and FcγRIIA, which mediate non-neutralizing antibody responses, was enhanced in non-transmitting mother-infant dyads and strongly correlated with ADCP. These findings suggest that Fc effector functions including ADCP protect against placental HCMV transmission. Taken together, our data indicate that future active and passive immunization strategies to prevent congenital HCMV infection should target Fc-mediated non-neutralizing antibody responses.

## Introduction

Human cytomegalovirus (HCMV) is the most common congenital infection worldwide, affecting 1 out of 150 births or nearly 1 million newborns annually (1, 2). Most congenital HCMV (cCMV) infections are asymptomatic, yet serious disease outcomes occur in 15-20% of cases including intrauterine growth restriction, neonatal multi-organ disease, neurodevelopmental impairment, and sensorineural hearing loss. Moreover, cCMV has been linked to an elevated risk of acute lymphoblastic leukemia (3–5) and is a leading infectious cause of stillbirth and neonatal death worldwide (6, 7). Regrettably, newborn screening for and public awareness of cCMV remains limited, leaving most cases undiagnosed and the true burden of disease underestimated (3, 8, 9). Despite the associated morbidity and mortality, there are no licensed vaccines or therapeutics to prevent cCMV. Thus, an improved understanding of the immune correlates of protection against congenital HCMV transmission is urgently needed to guide maternal active and passive immunization strategies.

HCMV is a host-restricted and ubiquitous β-herpesvirus with multiple envelope glycoproteins and complexes including glycoprotein B (gB) and gHgL “dimer,” which can associate with gO to form the gHgLgO “trimer” or pUL128/130/131 to form the “pentamer” complex (10). HCMV glycoproteins mediate viral entry, and following primary infection, the host remains latently infected for life (10–12). Globally, over 80% of reproductive-aged women are latently infected with HCMV, and congenital transmission occurs in maternal primary and nonprimary infection (i.e., reactivation from latency or reinfection with new strains) (2, 12–16). Mothers with primary infection have a 30% risk of placental transmission whereas those with nonprimary infection have a 1-4% risk (2, 17–19), indicating that preexisting maternal immunity partially protects against cCMV. In maternal primary infection, high avidity HCMV-specific IgG and anti-pentamer IgG binding are correlated with protection against transmission (20–23). Yet, maternal correlates of protection in nonprimary infection remain unclear, as HCMV glycoprotein-specific IgG binding and neutralizing antibody titers do not reliably correlate with protection against congenital transmission (24–26). Since 70-80% of cCMV infections occur in the setting of preexisting maternal immunity, understanding protective immune responses against *in utero* transmission in this population remains essential to preventing cCMV infection worldwide (2, 12, 16).

Identifying antibody responses that protect against placental HCMV transmission is necessary to develop novel vaccines and passive immunization strategies. Maternal treatment with HCMV hyperimmunoglobulin, a pooled preparation of high avidity, neutralizing HCMV-specific IgG, to prevent cCMV following primary infection has shown variable efficacy (27–32). Thus, a vaccine that will prevent maternal primary infection and/or placental transmission to the fetus remains a top public health priority. High avidity gB- and pentamer-specific IgG binding and neutralizing antibodies have been the main targets of HCMV vaccine development (10, 33); however, emerging evidence from vaccine trials indicates that non-neutralizing antibody functions may be key to preventing virus acquisition (34–37).

Antibodies can mediate polyfunctional responses including viral neutralization through the Fab region that binds antigen and effector functions through the Fc region that binds Fc receptors (FcRs) on innate immune cells. Whether non-neutralizing Fc-mediated antibody responses including antibody-dependent cellular phagocytosis (ADCP) and antibody-dependent cellular cytotoxicity (ADCC) protect against cCMV infection has not been explored to-date. In this study, we focused on ADCP since vaccine trials suggest that Fc effector functions independent of ADCC mediate protection against HCMV infection (34, 38). Moreover, we hypothesized that ADCP would be associated with protection against placental HCMV transmission since ADCP can eliminate virus:IgG immune complexes as well as virally-infected cells and because monocytes/macrophages expressing the FcγRs that mediate ADCP are enriched at the human maternal-fetal interface (39).

To identify immune correlates of protection against placental HCMV transmission, we measured anti- HCMV IgG specificities and anti-viral functions in maternal and cord blood sera from HCMV transmitting and non-transmitting mother-infant dyads identified as donors to a large U.S.-based public cord blood bank. In our primary analysis, we compared 13 predefined binding and functional anti-HCMV antibody responses in transmitting versus non-transmitting women. In an exploratory analysis, we applied a systems serology approach to define differences between transmitting and non-transmitting mother- infant dyads and quantified HCMV-specific IgG engagement of host FcγRs. Insights from this study of binding and functional antibody responses in HCMV transmitting and non-transmitting mother-infant dyads can inform vaccine and therapeutic development to prevent cCMV and ameliorate a major cause of pediatric morbidity worldwide.

## Results

### Baseline characteristics of HCMV transmitting and non-transmitting mother-infant dyads

Our study included sera from 81 mother-infant dyads recruited as donors to the Carolinas Cord Blood Bank (CCBB), a U.S.-based public cord blood bank with 130,000 collections and 45,000 banked donors to-date (**Supplementary Figure 1**). Donor mothers were screened for total anti-HCMV antibodies and if present, cord blood plasma was tested by qPCR for replicating HCMV. Cord blood units that contained replicating HCMV were excluded from banking for cord blood transplantation and were retained for research. Congenital HCMV infection (cCMV) was defined based on the presence of HCMV viremia in the donated cord blood plasma. Forty-one dyads with congenital HCMV infection (“HCMV transmitting”) were matched to forty dyads with HCMV IgG seropositive mothers that gave birth to cCMV uninfected infants (“HCMV non-transmitting”). Matching criteria included infant sex, infant race, maternal age (+/- 3 years), and delivery year (+/- 3 years). Baseline demographic and clinical characteristics were comparable between HCMV transmitting and non-transmitting dyads, though transmitting dyads had a non-significantly higher rate of Cesarean section (56% vs. 40%, Fisher’s exact p = 0.22, **Table 1**). The majority of identified cCMV cases were male (58.5%), higher than the proportion of males in the cord blood bank overall (51.4%), which may suggest an increased susceptibility to infection for male fetuses, though studies on sex differences in cCMV infection are lacking (40).

**Table 1.** Baseline characteristics of HCMV transmitting and non-transmitting cord blood bank donor mother-infant dyads.

| Mother-infant dyad characteristics | HCMV transmitting (n=41) | HCMV non-transmitting (n=40) |
| --- | --- | --- |
| **Infant sex, n (%)** |  |  |
| Female | 17 (41.5%) | 16 (40.0%) |
| Male | 24 (58.5%) | 24 (60.0%) |
| **Infant race/ethnicity, n (%)** |  |  |
| Caucasian | 26 (63.4%) | 24 (60.0%) |
| African American | 8 (19.5%) | 8 (20.0%) |
| Hispanic | 2 (4.9%) | 2 (5.0%) |
| Other | 5 (12.2%) | 6 (15.0%) |
| **Maternal age (years), median [IQR]** | 27 [23, 31] | 28 [24, 33] |
| **Gestational age (months), median [IQR]** | 39.0 [39.0, 40.0] | 39.0 [38.0, 40.0] |
| **Delivery year, median [range]** | 2013 [2010, 2015] | 2012 [2008, 2017] |
| **Delivery type, n (%)** |  |  |
| Vaginal | 18 (43.9%) | 24 (60.0%) |
| Cesarean section | 23 (56.1%) | 16 (40.0%) |
| **Maternal HCMV IgG avidity index, median [IQR]** | 77.6 [70.4-84.5] | 79.5 [73.3-86.3] |
| **Maternal HCMV IgM seropositivity, n (%)** |  |  |
| Seropositive | 11 (26.8%) | 2 (5.0%) |
| Seronegative | 30 (73.2%) | 38 (95.0%) |
| **Maternal sera HCMV DNAemia** |  |  |
| Positive | 11 (26.8%) | 15 (37.5%) |
| Negative | 30 (73.2%) | 25 (62.5%) |
| **Maternal sera HCMV viral copies, median [range]<sup>a</sup>** | 346 [256-1052] | 365 [260-719] |
| **Cord blood sera HCMV viral copies, median [range]<sup>c</sup>** | 727 [137-18,100] | ND |

ND = not detected
<sup>a</sup> Maternal HCMV viral copies listed in viral copies/mL, lower limit of detection = 250 copies/mL
<sup>b</sup> Cord blood HCMV viral copies detected listed in IU/mL, lower limit of detection = 137 copies/mL
HCMV transmitting and non-transmitting dyads were matched on maternal age (+/- 3 years), infant race, sex, and delivery year (+/- 3 years)

To assess whether mothers had primary or nonprimary HCMV infection during pregnancy, we measured anti-HCMV IgG avidity and IgM responses (41). Whole virion HCMV IgG avidity indexes were comparable between HCMV transmitting (median [range]: 77.6 [46-109]) and non-transmitting (median [range]: 79.5 [65-100]) mothers. However, 11/41 (26.8%) of HCMV transmitting mothers had detectable HCMV-specific IgM responses compared to only 2/40 (5%) of non-transmitting mothers (**Table 1**). Thus, while the majority of mothers in both groups had high avidity IgG responses and no detectable IgM responses at the delivery timepoint, a higher proportion of mothers in the transmitting group had detectable HCMV-specific IgM, which could indicate a higher rate of primary infection or reinfection during pregnancy in the transmitting group (41, 42). To further assess comparability between groups, we quantified HCMV viral loads in maternal sera and found that a similar proportion of transmitting (26.8%) and non-transmitting (37.5%) mothers had detectable low-level HCMV DNAemia (median [range] in copies/mL: 346 [256-1052] in transmitting vs. 365 [260-719] in non-transmitting; **Table 1**), consistent with previous observations in healthy HCMV seropositive women (43).

### Maternal and cord blood sera from HCMV transmitting dyads have high levels of anti-HCMV IgG

We previously observed that HCMV-specific IgG transfer across the placenta may be compromised in cCMV infection, a phenomenon primarily related to maternal hypergammaglobulinemia in HCMV transmitting women (44). Thus, we first measured placental HCMV-specific IgG transfer in transmitting and non-transmitting mother-infant dyads. We quantified IgG binding against 3 distinct HCMV strains including TB40E (an endotheliotropic strain expressing pentamer), AD169r (a lab-adapted strain with repaired pentamer expression), and Toledo (a low-passage clinical isolate lacking pentamer). Within mother-infant pairs, HCMV-specific IgG binding was lower in cord blood compared to maternal sera in transmitting but not non-transmitting dyads (**Figure 1A**). However, placental IgG transfer efficiency was only slightly lower in transmitting compared to non-transmitting dyads (**Figure 1B**). HCMV transmitting mothers also had higher total IgG levels (i.e., hypergammaglobulinemia), which negatively correlated with HCMV-specific IgG transfer efficiency (**Supplementary Figure 2A-B**). Despite slightly decreased placental IgG transfer ratios, whole virion HCMV-specific IgG titers were comparable in the cord blood of infected and uninfected infants (**Figure 1C**). In contrast, HCMV glycoprotein-specific IgG titers were significantly elevated (2.5-10 fold) in transmitting compared to non-transmitting dyads (**Figure 1D-E**). When adjusting for total IgG levels, non-transmitting women demonstrated higher relative binding to HCMV virions and cell-associated native gB, while IgG binding against pentamer, gHgLgO, and gHgL remained significantly higher in transmitting women (**Supplementary Figure 2C-D).**

**Figure 1.**
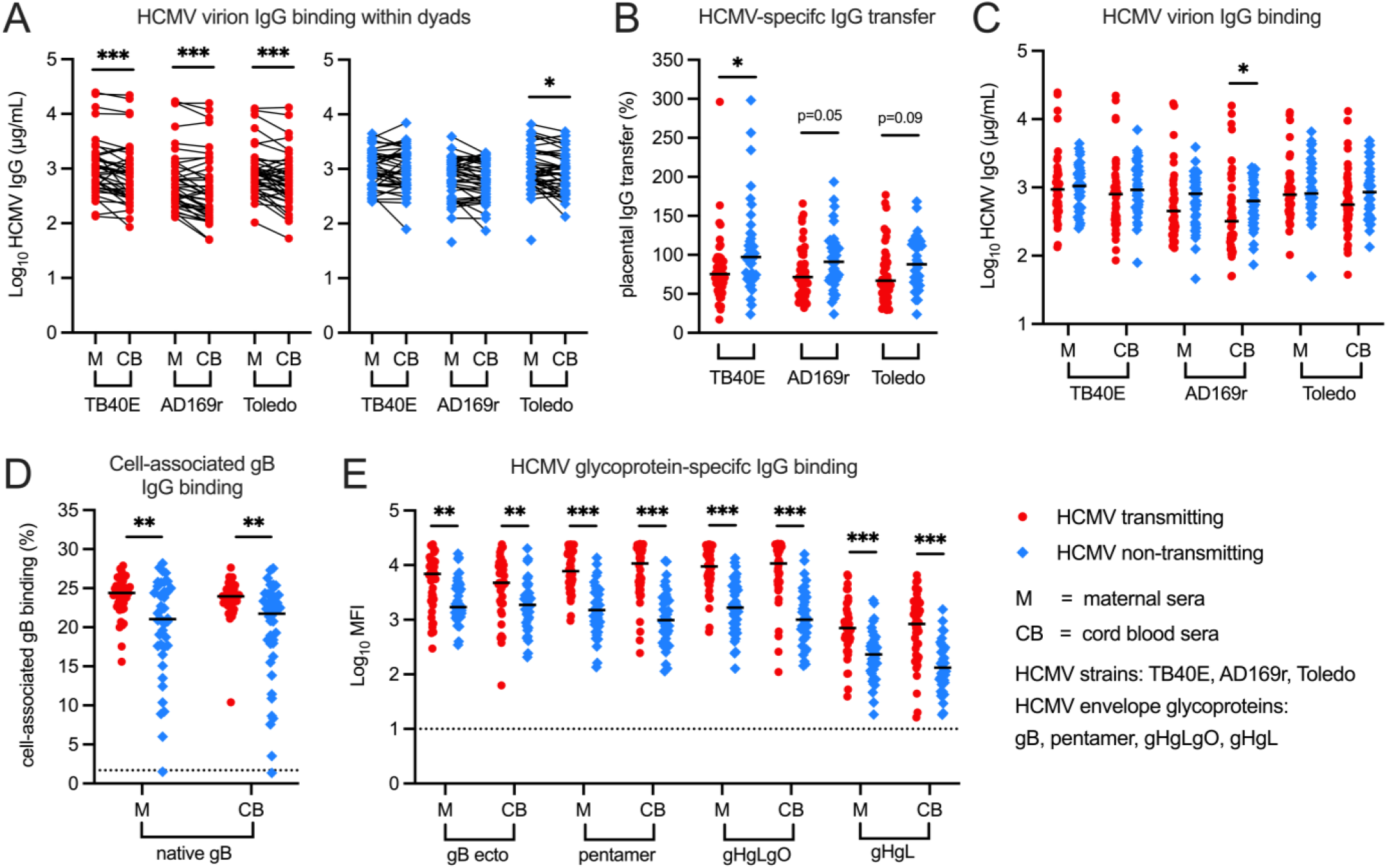
Maternal and cord blood sera from HCMV transmitting dyads have high levels of HCMV-specific IgG despite moderately reduced transplacental IgG transfer compared to non- transmitting dyads. HCMV-specific IgG levels against HCMV strains TB40E, AD169r, and Toledo were measured using whole virion enzyme-linked immunosorbent assay (ELISA). Cell-associated gB IgG binding was quantified using a flow-based assay with HEK293T cells transfected with full-length gB and HCMV glycoprotein-specific IgG binding was measured using a Luminex-based binding antibody multiplex assay. IgG binding responses in maternal (M) and cord blood (CB) sera were compared between and within HCMV transmitting (red circles, n = 41) and non-transmitting (blue squares, n = 40) mother-infant dyads. (A) Whole virion HCMV-specific IgG levels in paired maternal and cord blood sera. (B) Transplacental transfer efficiency of HCMV-specific IgG calculated as (cord blood HCMV-specific IgG concentration)/(maternal HCMV-specific IgG concentration)x100%. (C) Whole virion HCMV-specific IgG levels in transmitting versus non-transmitting dyads. (D) IgG binding to cell-associated gB via flow cytometry. (E) Mean fluorescence intensity (MFI) proportional to IgG binding to HCMV glycoproteins. gB ecto = gB ectodomain. Dotted lines indicate the lower limit of detection. Black bars denote median. FDR-corrected P values reported for Mann-Whitney *U* test or Wilcoxon signed-rank test. * P < 0.05, ** P < 0.01, *** P < 0.001.

### Low avidity HCMV-specific IgG is enriched in the cord blood of cCMV-infected infants

Transplacental transfer of low-avidity HCMV-specific IgG complexed to HCMV virions has been posited to increase HCMV transmission risk (45) and we previously observed that low avidity HCMV-specific IgG may be preferentially transferred across the placenta in HCMV transmitting pregnancies (44). Thus, we next measured HCMV-specific IgG avidity in paired maternal and cord blood samples. Maternal sera from non-transmitting pairs had higher avidity IgG binding to AD169r and Toledo strains, but not TB40E, compared to transmitting dyads (**Figure 2A**). Within mother-infant pairs, whole virion HCMV IgG binding avidity was significantly lower in the cord blood compared to maternal sera in the transmitting but not the non-transmitting groups (**Figure 2B**). HCMV glycoprotein-specific IgG avidity was also lower in the cord blood of infected compared to uninfected infants (**Figure 2C**), though no differences in glycoprotein-specific IgG avidity were observed within paired sera from mother-infant dyads. In a sensitivity analysis excluding mothers with HCMV-specific IgM responses as a surrogate biomarker for recent maternal primary infection or reinfection, differences in whole virion HCMV IgG binding avidity persisted but most differences in HCMV glycoprotein-specific IgG binding avidity were no longer statistically significant (**Supplementary Figure 3A-C**).

**Figure 2.**
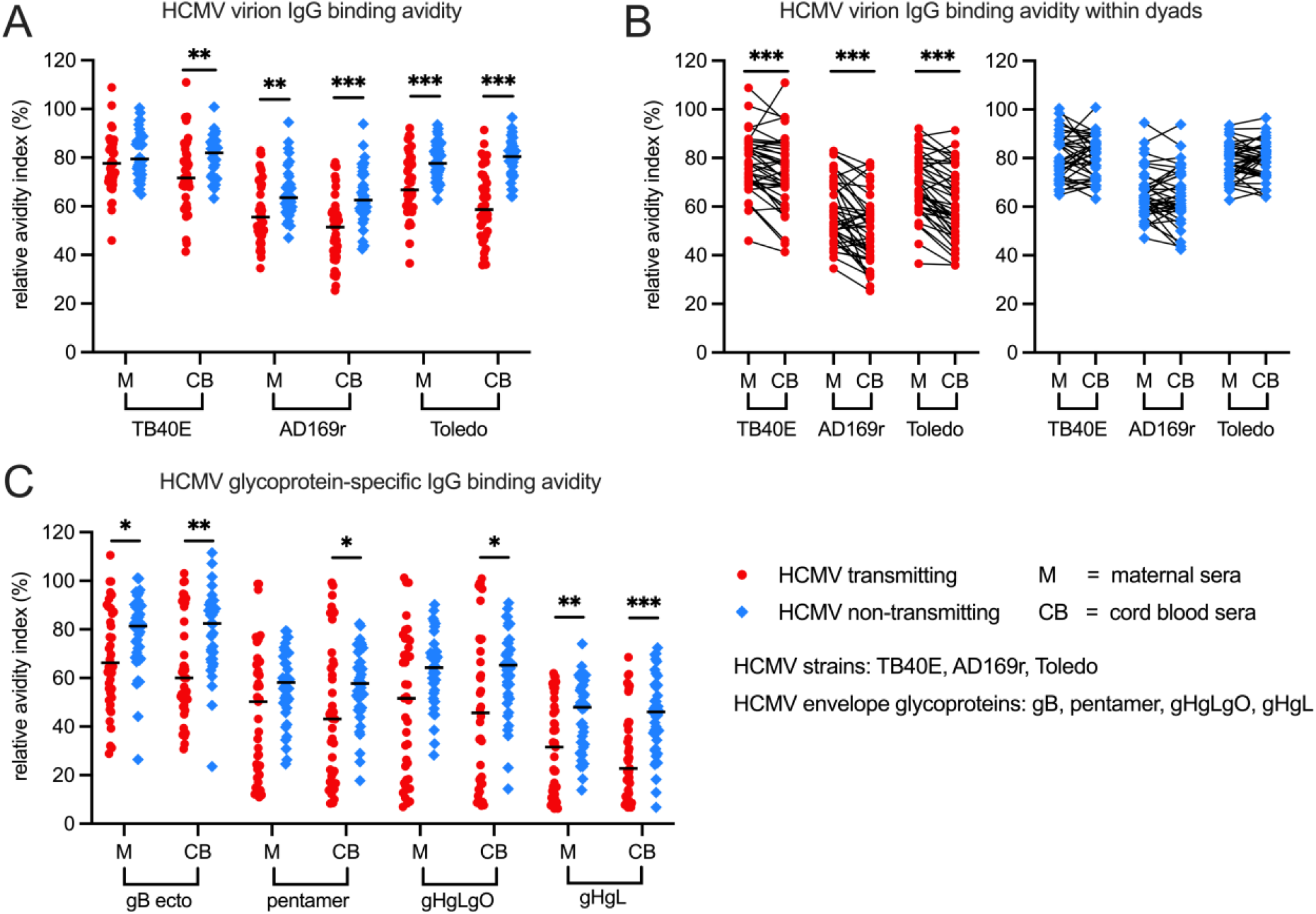
Low avidity HCMV-specific IgG is enriched in the cord blood of cCMV-infected infants. HCMV-specific IgG binding avidity against HCMV strains TB40E, AD169r, and Toledo were measured using whole virion enzyme-linked immunosorbent assay (ELISA) with an additional dissociation step using urea and relative avidity index (RAI) was calculated as (OD with urea/OD without urea)x100%. HCMV glycoprotein-specific IgG binding avidity was measured using a Luminex-based binding antibody multiplex assay with an additional dissociation step with sodium citrate and RAI was calculated as (MFI with sodium citrate/MFI with 1X PBS)x100%. IgG binding avidity in maternal (M) and cord blood (CB) sera were compared between and within HCMV transmitting (red circles, n = 41) and non-transmitting (blue squares, n = 40) mother-infant dyads. (A) Whole virion HCMV-specific IgG binding avidities in transmitting versus non-transmitting dyads. (B) Whole virion HCMV-specific IgG binding avidities in paired maternal and cord blood sera. (C) HCMV glycoprotein-specific IgG binding avidities in transmitting versus non-transmitting dyads. gB ecto = gB ectodomain. Black bars denote median. FDR- corrected P values reported for Mann-Whitney *U* test or Wilcoxon signed-rank test. * P < 0.05, ** P < 0.01, *** P < 0.001.

### Neutralizing and non-neutralizing antibody responses differ in HCMV transmitting and non- transmitting dyads

In addition to measuring HCMV-specific IgG binding magnitude and avidity, we quantified anti-viral functions of maternal and cord blood sera from transmitting and non-transmitting dyads. Unexpectedly, HCMV neutralizing titers were approximately 1.5-4 fold higher in transmitting versus non-transmitting dyads across HCMV strains and cell types (**Figure 3A-D**). Within mother-infant dyads, neutralizing titers were similar in the maternal compared to cord blood sera in both groups. In contrast, ADCP, an Fc-mediated non-neutralizing antibody response, against HCMV was higher in non-transmitting compared to transmitting mothers (**Figure 3E**). This difference was statistically significant for the Toledo HCMV strain (p = 0.0057) and remained significant after correction for multiple comparisons (p

**Figure 3.**
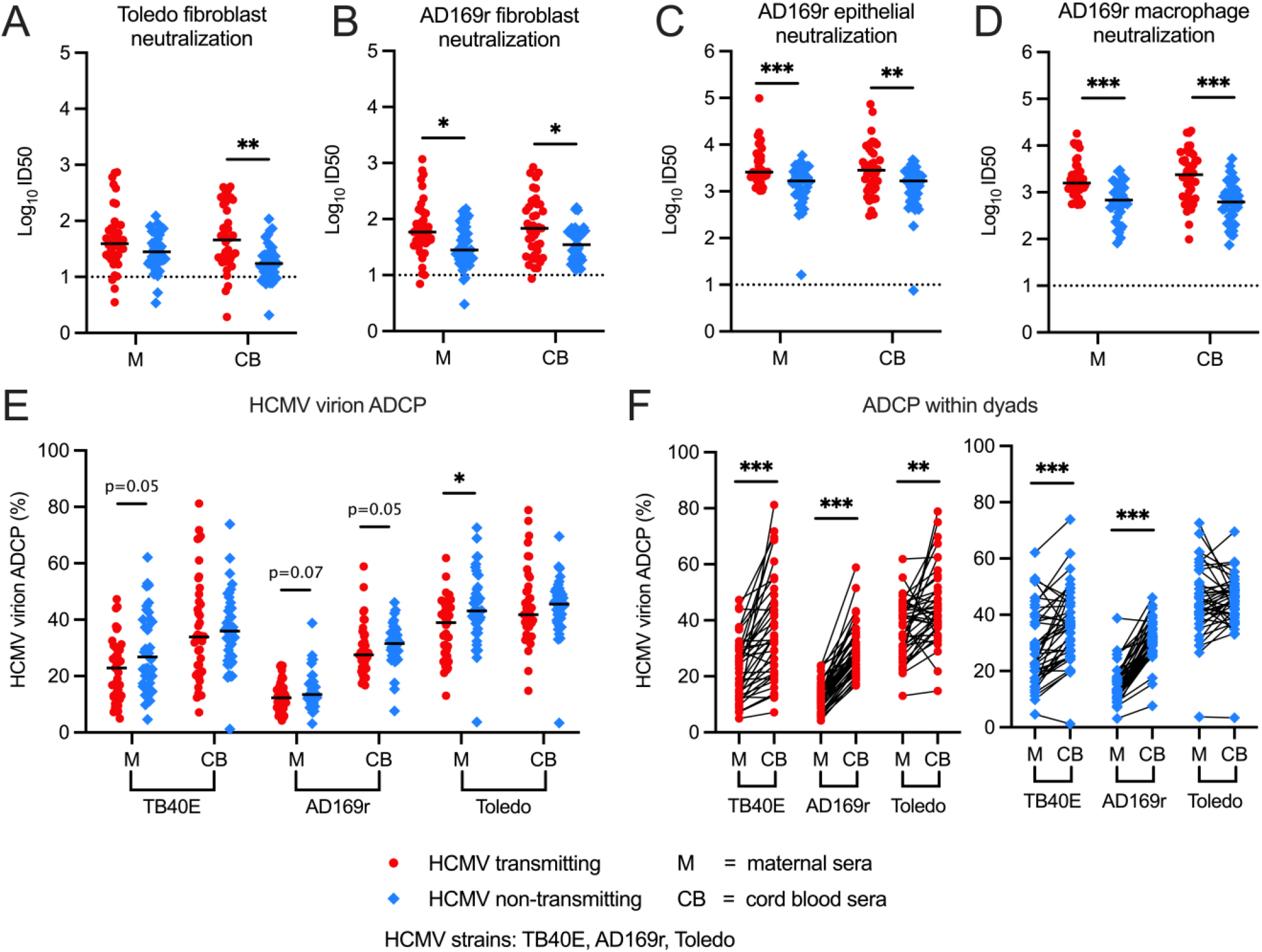
Higher non-neutralizing HCMV-specific ADCP responses and lower neutralizing responses in HCMV non-transmitting compared to transmitting mother-infant dyads. Functional anti-viral antibody responses in maternal (M) and cord blood (CB) sera were compared between and within HCMV transmitting (red circles, n = 41) and non-transmitting (blue squares, n = 40) mother-infant dyads. Neutralization was measured by HCMV intermediate early gene 1 (IE1) staining and antibody titers were calculated as the inhibitory dilution 50 (ID50), equivalent to the sera dilution that inhibited 50% of the max infection in virus only wells. Neutralization titers against (A) HCMV Toledo strain in fibroblasts (HFFs) and (B-D) HCMV AD169r strain in (B) fibroblasts (HFFs), (C) epithelial cells (ARPEs) and (D) differentiated macrophages (THP-1s treated with PMA). (E-F) Antibody-dependent cellular phagocytosis (ADCP) of AF647 fluorophore-conjugated HCMV virions by THP1 monocytes was quantified using a flow-based assay and calculated as percentage AF647 positive cells. (E) HCMV-specific ADCP responses in transmitting versus non-transmitting dyads. (F) HCMV-specific ADCP responses in paired maternal and cord blood sera. Black bars denote median. Dotted lines for neutralization assays indicate the starting dilution corresponding to the lower limit of detection (ID50 = 10). FDR-corrected P values reported for Mann-Whitney *U* test or Wilcoxon signed-rank test. * P < 0.05, ** P < 0.01, *** P < 0.001.

= 0.011) with a trend towards statistical significance for TB40E (p = 0.053) and AD169r (p = 0.068). Interestingly, within mother-infant pairs, whole virion HCMV ADCP responses were highly enriched in the cord blood sera compared to maternal sera within both transmitting and non-transmitting groups (**Figure 3F**), suggesting preferential transfer of ADCP-mediating IgG across the placenta. The enhanced transfer of ADCC-eliciting antibodies across the placenta has been described in healthy pregnancies (46), yet our results indicate that IgG capable of diverse Fc effector functions such as ADCP are also selectively transferred from mother-to-fetus.

### Maternal ADCP and high-avidity glycoprotein-specific IgG binding correlate with decreased risk of congenital HCMV infection

For our primary immune correlate analysis, we predefined 13 maternal antibody responses (**Table 2**) that we hypothesized would be associated with decreased risk of placental HCMV transmission. Using univariate logistic regression, 12 out of the 13 predefined variables were significantly (p < 0.05) associated with our primary endpoint of congenital HCMV infection. Maternal HCMV glycoprotein- specific IgG binding magnitude and neutralization titers were associated with increased risk of congenital transmission whereas IgG binding avidity and ADCP were associated with decreased transmission risk (**Table 2**). HCMV glycoprotein-specific IgG binding magnitude and neutralization titers remained significantly associated with higher risk of transmission after adjusting for maternal total IgG levels and maternal HCMV-specific IgM status; however, glycoprotein-specific IgG binding avidity was no longer statistically significant after adjusting for these variables (**Supplementary Table 1**). ADCP against Toledo, an HCMV strain lacking pentamer, remained significantly associated with protection against transmission status in both adjusted univariate regression models (**Supplementary Table 1**).

**Table 2.** Univariate and LASSO regression analysis of maternal humoral immune correlates of congenital HCMV transmission. Logistic regression analysis on 13 primary variables of maternal sera anti-HCMV antibody responses comparing HCMV transmitting and HCMV IgG seropositive non-transmitting mother-infant dyads.

|  | Univariate logistic regression |  | LASSO regression <sup>c</sup> |
| --- | --- | --- | --- |
| Antibody response | Beta coefficient (SE)<sup>b</sup> | P value | Beta coefficient <sup>b</sup> |
| IgG binding to cell-associated gB (%) | 0.206 (0.068) | **0.003** | - |
| gB ectodomain IgG binding (Log MFI) | 0.738 (0.229) | **0.001** | - |
| pentamer IgG binding (Log MFI) | 1.749 (0.384) | **<0.0001** | 1.60 |
| gHgLgO IgG binding (Log MFI) | 1.469 (0.323) | **<0.0001** | - |
| gB IgG avidity (%) | -0.030 (0.013) | **0.021** | -0.33 |
| pentamer IgG avidity (%) | -0.023 (0.011) | **0.033** | - |
| gHgLgO IgG avidity (%) | -0.027 (0.011) | **0.011** | -0.23 |
| Fibroblast neutralization (Log ID50)<sup>a</sup> | 0.688 (0.258) | **0.008** | - |
| Epithelial neutralization (Log ID50)<sup>a</sup> | 1.543 (0.424) | **<0.0001** | - |
| Macrophage neutralization (Log ID50)<sup>a</sup> | 1.544 (0.397) | **<0.0001** | - |
| Whole virion Toledo ADCP (%) | -0.058 (0.022) | **0.008** | -0.14 |
| Whole virion TB40E ADCP (%) | -0.040 (0.019) | **0.032** | - |
| Whole virion AD169r ADCP (%) | -0.077 (0.043) | 0.073 | - |

<sup>a</sup> Neutralization measured against HCMV strain AD169r
<sup>b</sup> Positive beta coefficients are associated with increased risk and negative beta coefficients are associated with decreased risk of placental HCMV transmission
<sup>c</sup> Beta coefficients shown for significant variables included in model after LASSO feature selection
“-” Symbol indicates that the variable was not selected in the LASSO regression model
ADCP = antibody-dependent cellular phagocytosis
SE = standard error
**Bold indicates statistical significance (P value < 0.05)**

For our multivariable immune correlate analysis, we used least absolute shrinkage and selection operator (LASSO) modeling. Many immune parameters included in our predefined primary analysis were strongly correlated with each other (**Supplementary Figure 4**). Due to this high degree of multicollinearity, LASSO was employed for feature selection. LASSO is an approach to minimize overfitting a regression model that can shrink the coefficients of poorly predictive variables to zero, thereby removing them from the model. First, the cohort was randomly split into a training and test dataset and a k-fold cross-validation approach was employed to train the LASSO model. LASSO- selected features included magnitude of IgG binding to pentamer, avidity of gB IgG binding, avidity of gHgLgO IgG binding and ADCP against Toledo strain (**Table 2**). Notably, this 4-parameter LASSO model had a 0.75 accuracy (95% CI: 0.48-0.93) in predicting risk of congenital HCMV transmission (with 1.00 equaling perfect predictive capacity and 0.50 equaling the random prediction rate with no information). Consistent with the univariate analyses, the magnitude of pentamer IgG binding was associated with increased risk of congenital transmission whereas glycoprotein-specific IgG binding avidity and ADCP were associated with decreased risk of congenital transmission.

### HCMV glycoprotein-specific IgG binding to Fc receptors differs in HCMV transmitting and non-transmitting dyads

In an exploratory analysis, we quantified FcR engagement by HCMV-specific IgG in maternal and cord blood sera to examine the role of Fc-mediated antibody responses and placental IgG transfer in congenital HCMV transmission. Since FcRs can transfer immune complexes across the placenta, we explored if differential HCMV-specific IgG binding to FcRs in maternal versus cord blood sera was associated with placental transmission (47). Using a multiplex FcR binding assay, we measured HCMV glycoprotein-specific IgG binding to the neonatal Fc receptor (FcRn), which regulates IgG half-life and placental IgG transfer, and the Fcγ receptors FcγRI, FcγRIIA, FcγRIIB and FcγRIIIA, which mediate Fc effector functions such as ADCP (47–49). HCMV-specific IgG binding to FcRs was significantly higher in transmitting compared to non-transmitting dyads across most antigens (**Supplementary Figure 5A-G**). Since transmitting dyads had higher binding to gB-, pentamer-, gHgLgO- and gHgL-coated multiplex beads at baseline (**Figure 1E**), we normalized FcR binding to total antigen-specific IgG binding for each HCMV glycoprotein measured. After normalization, transmitting and non-transmitting dyads had similar magnitude HCMV-specific IgG binding to FcRn, FcγRIIB and FcγRIIIA between and within mother-infant pairs (**Supplementary Figure 6A, C-E**). However, HCMV glycoprotein-specific IgG binding to FcγRI was significantly higher in non-transmitting compared to transmitting dyads (**Figure 4A**). Within mother-infant pairs, HCMV glycoprotein-specific IgG binding to FcγRI was also highly enriched in the cord blood of uninfected infants (**Figure 4B**). HCMV viral loads in the cord blood of infected infants were significantly negatively correlated with FcγRI binding to maternal gHgLgO- (Spearman ρ = -0.3, p = 0.019), pentamer- (Spearman ρ = -0.36, p = 0.020), and gB- (Spearman ρ = -0.32, p = 0.037) specific IgG, indicating that HCMV-specific IgG engagement of FcγRI may help control HCMV viremia. Conversely, glycoprotein-specific IgG binding to FcγRIIA was enhanced in the cord blood of infected infants with preferential transfer across the placenta within mother-infant dyads, though this was only observed for the high-affinity variant FcγRIIAH131 (**Figure 4C-D, Supplementary Figure 6B**).

**Figure 4.**
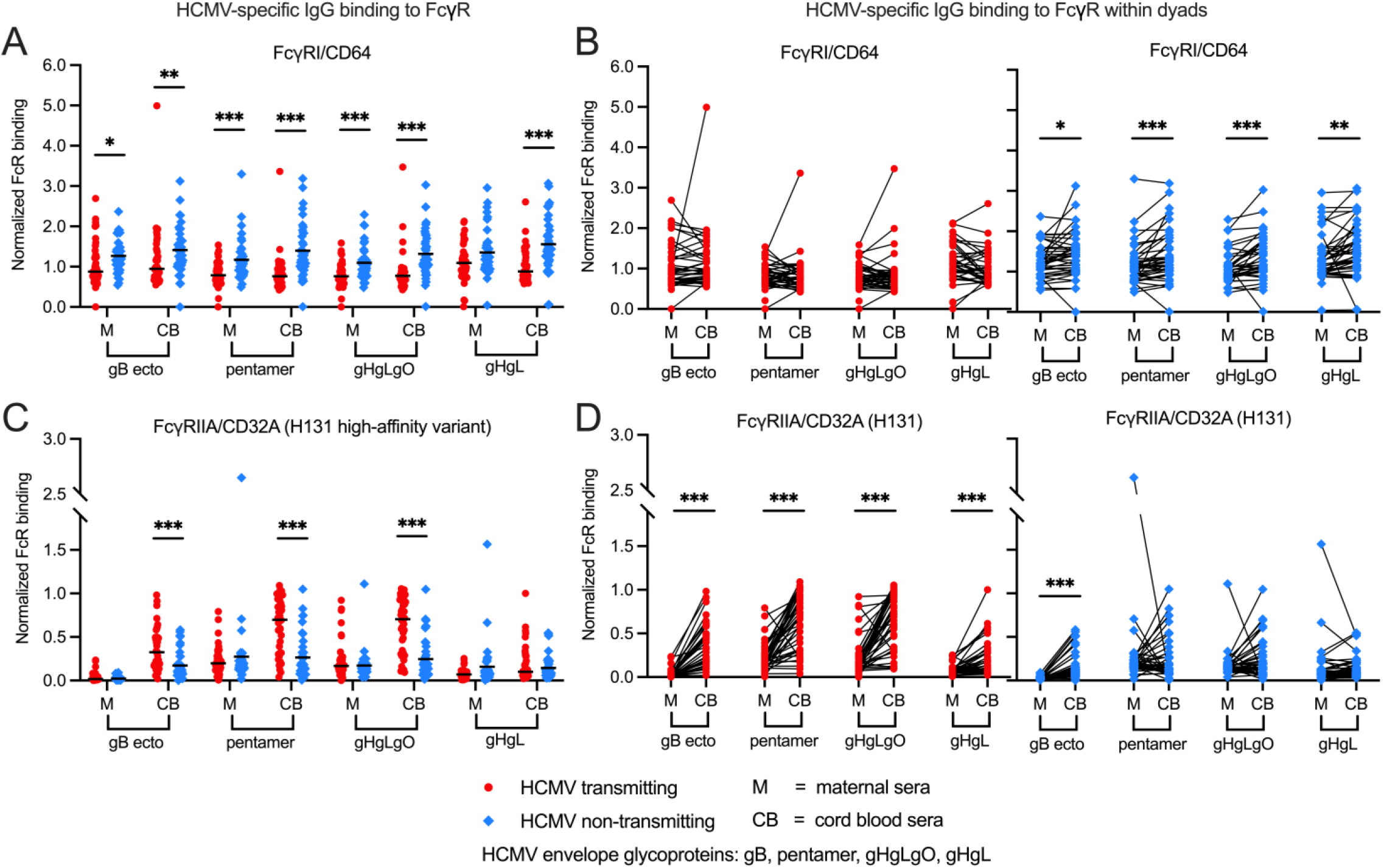
HCMV glycoprotein-specific IgG binding to FcγRI and FcγRIIA differs in HCMV transmitting and non-transmitting dyads. HCMV glycoprotein-specific IgG binding to Fcγ receptors (FcγRs) was measured using a Luminex-based binding antibody multiplex assay with a biotinylated FcγR and streptavidin-PE detection antibody. Normalized antigen-specific IgG binding to host FcγRs was compared between and within HCMV transmitting (red circles, n = 41) and non-transmitting (blue squares, n = 40) mother-infant dyads. (A-B) HCMV-specific IgG binding to FcγRI in (A) transmitting versus non-transmitting dyads and (B) paired maternal and cord blood sera. (C-D) HCMV-specific IgG binding to FcγRIIA high affinity H131 variant in (C) transmitting versus non-transmitting dyads and (D) paired maternal and cord blood sera. gB ecto = gB ectodomain. Black bars denote median. FDR- corrected P values reported for Mann-Whitney *U* test or Wilcoxon signed-rank test. * P < 0.05, ** P < 0.01, *** P < 0.001.

### Systems serology analysis reveals distinct anti-HCMV antibody profile in HCMV transmitting compared to non-transmitting dyads

We next employed a principal components analysis (PCA) across all 50 humoral immune variables tested to define differences in HCMV-specific antibody responses in transmitting versus non- transmitting dyads. In the maternal and cord blood sera, PC1 accounted for 57 and 59% of the variance, respectively; however, PC2, which accounted for 16 and 17% of the variance, was superior at delineating between transmitting and non-transmitting dyads (**Figure 5A-B**). The top 12 contributors to PC2 were the same in the PCA of maternal and cord blood sera and included HCMV-specific IgG binding avidity, IgG binding magnitude, and ADCP (**Figure 5C-D**). These PCA results demonstrate that high avidity HCMV-specific IgG and ADCP responses were enriched in the maternal and cord blood sera of non-transmitting compared to transmitting dyads, further validating the findings in our primary analysis.

**Figure 5.**
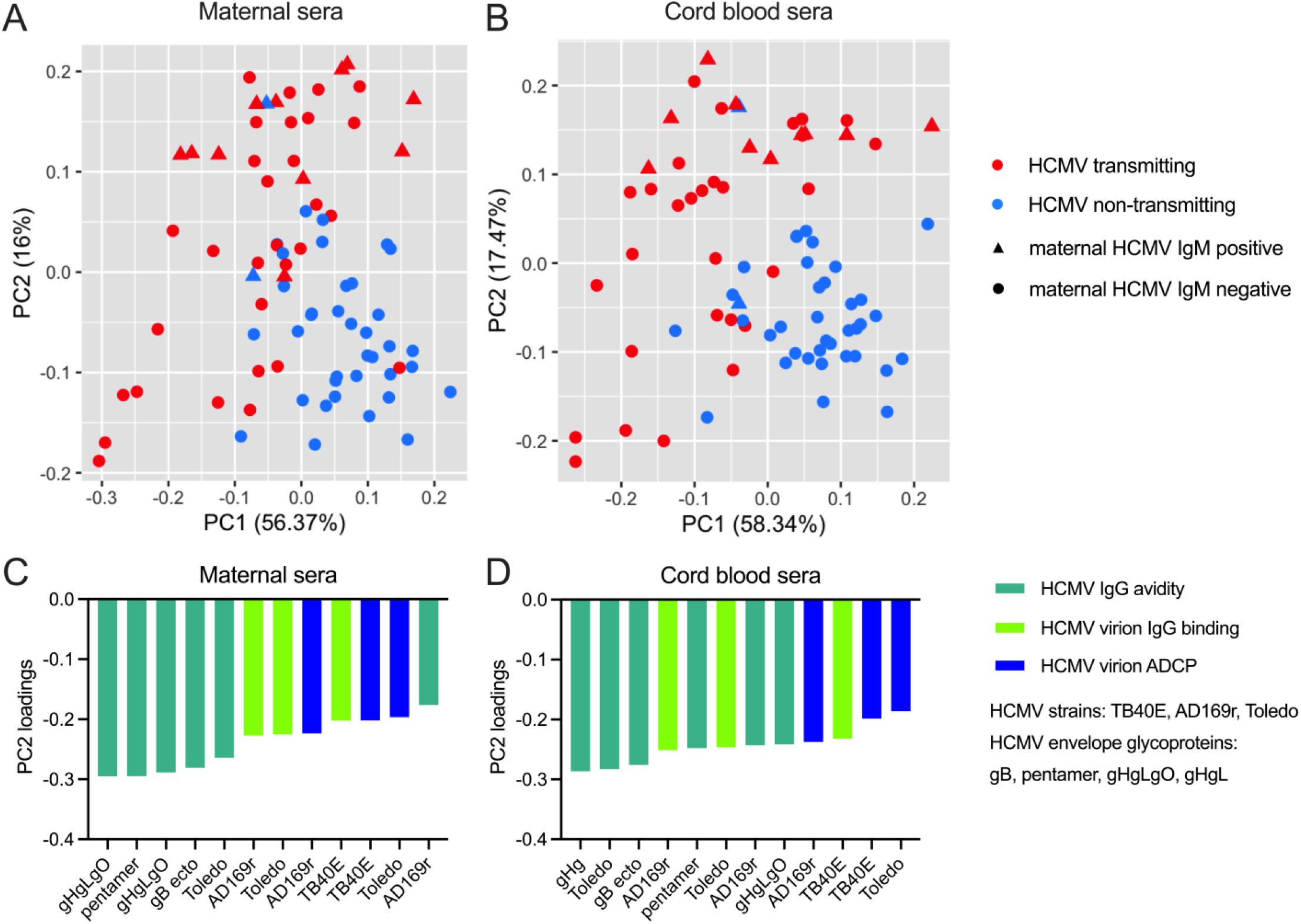
Principal components analysis (PCA) reveals distinct HCMV-specific antibody responses in HCMV transmitting compared to non-transmitting dyads. Principal components analysis (PCA) across all 50 antibody responses measured for HCMV transmitting (red, n = 41) and non-transmitting (blue, n = 40) mother-infant dyads. Triangles (n=14) indicate dyads where mothers screened positive for HCMV-specific IgM responses and circles (n=67) indicate dyads where mothers had no detectable HCMV-specific IgM responses. Scatterplot of PC1 and PC2 for (A) maternal and (B) cord blood sera. Bar plot shows the magnitude of the top twelve antibody responses contributing to PC2 in (C) maternal and (D) cord blood sera.

### Maternal HCMV-specific IgG signaling through FcγRI and FcγRIIA is associated with protection against congenital HCMV infection

Since our predefined immune correlate and exploratory analyses both implicated Fc-mediated effector functions as protective against congenital HCMV transmission, we sought to further validate this novel finding by using a cell-culture based assay to evaluate anti-viral IgG activation of host FcγRs (50, 51). This approach leverages mouse thymoma BW cell lines stably transfected to express chimeric FcγRs with an extracellular human FcγR domain fused to an intracellular mouse CD3ζ signaling domain (**Figure 6A**). Using this previously validated model system with mouse IL-2 as a quantitative read-out for FcγR activation, we quantified HCMV-specific IgG signaling through FcγRI and FcγRIIA. We measured HCMV-specific IgG signaling through FcγRI and FcγRIIA because 1) there were differences in IgG binding to these FcγRs in transmitting versus non-transmitting dyads (**Figure 4**) and 2) FcγRI and FcγRIIA mediate ADCP, which we identified as associated with protection against congenital HCMV transmission. We first used flow cytometry to confirm that each previously validated cell line was exclusively expressing the FcγR of interest (**Figure 6B**). HCMV-specific IgG signaling through FcγRI was 3-4 fold higher when reporter cells were stimulated with maternal or cord blood sera from non-transmitting versus transmitting dyads (**Figure 6C**). HCMV-specific IgG signaling through FcγRIIA was similar in transmitting and non-transmitting mothers, yet there was a trend towards significantly increased FcγRIIA activation by HCMV-specific IgG in cord blood from non-transmitting pairs (**Figure 6D**). Moreover, a sensitivity analysis excluding outliers from both groups (ROUT method, Q = 1%) revealed that HCMV-specific IgG from non-transmitting dyads was significantly better at signaling through both FcγRI and FcγRIIA (**Supplementary Figure 7A-B**). Within mother-infant pairs, HCMV-specific IgG activation of FcγRI and FcγRIIA was significantly lower in cord blood versus maternal sera in both groups (**Figure 6E-F**). While HCMV glycoprotein-specific IgG binding to FcγRI and FcγRIIA correlated weakly with ADCP (**Supplementary Figure 4**), HCMV-specific IgG signaling through FcγRI and FcγRIIA correlated strongly with ADCP responses in maternal and cord blood sera (**Figure 6G-H**). These results imply that robust HCMV-specific IgG engagement of host FcγRI/IIA mediates effective monocyte/macrophage virion phagocytosis and protection against placental HCMV transmission.

**Figure 6.**
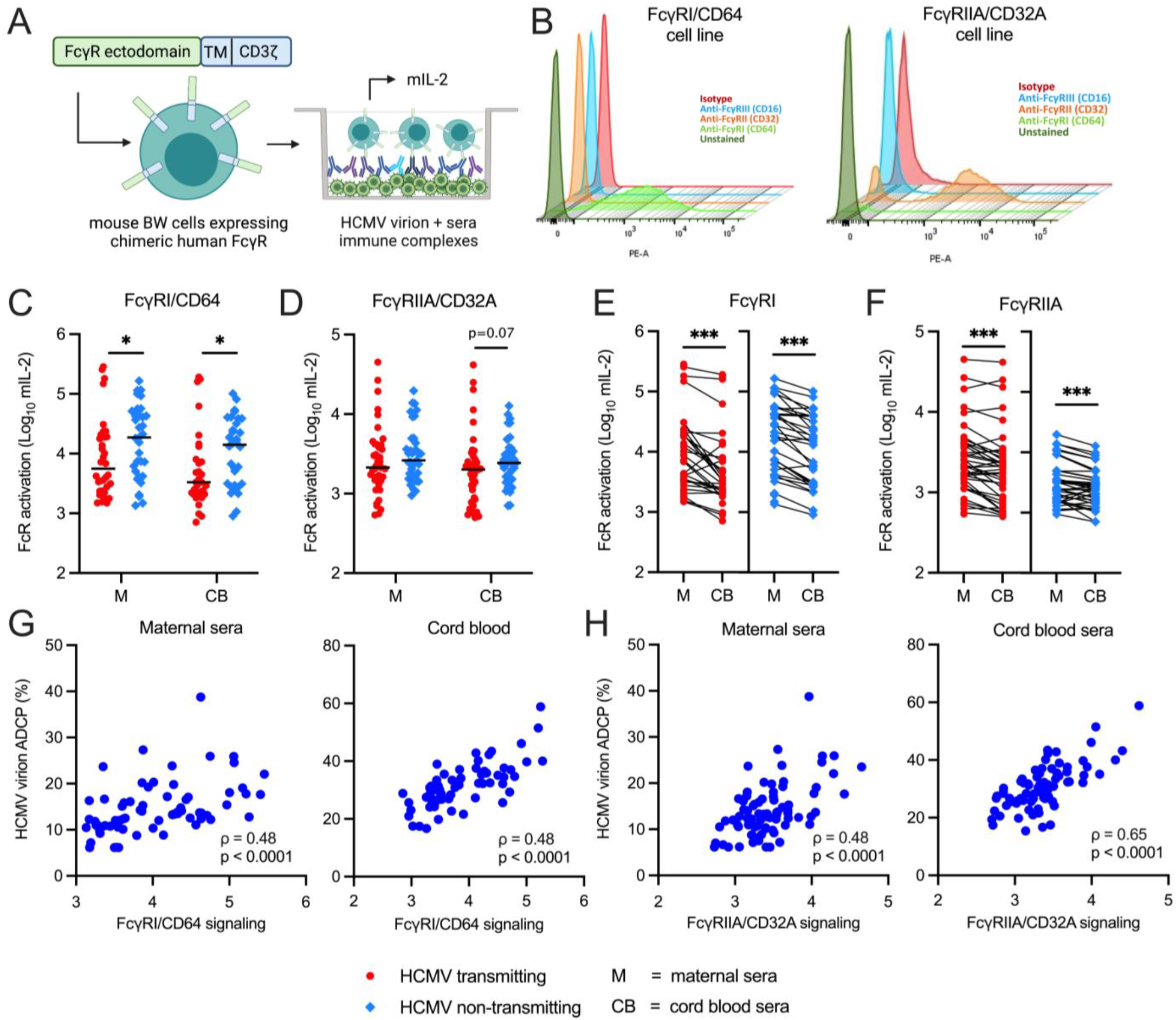
HCMV-specific IgG signaling through FcγRI and FcγRIIA is increased in HCMV non-transmitting compared to transmitting mother-infant dyads and strongly correlated with ADCP. HCMV-specific IgG activation of FcγRs was measured using maternal (M) and cord blood (CB) sera from HCMV transmitting (red circles, n = 41) and non-transmitting (blue squares, n = 40) mother-infant dyads. (A) To quantify HCMV-specific IgG activation of FcγRs, mouse BW cell lines stably expressing chimeric human FcγRs fused to a mouse CD3ζ signaling domain were co-cultivated with virus:sera immune complexes for 20 hours. Activation of FcγRs by immune complexes triggered CD3ζ signaling and mouse IL-2 (mIL-2) secretion, which was measured by ELISA as a quantitative read-out of HCMV-specific IgG signaling through host FcγRs. (B) Flow cytometry of BW cell lines stained with anti-FcγRI/CD64 (light green), anti-FcγRII/CD32 (orange), anti-FcγRIII/CD16 (blue) and isotype control (red) PE-conjugated antibodies to confirm FcγR expression. HCMV-specific IgG activation of (C) FcγRIA and (D) FcγRIIA in transmitting and non-transmitting dyads. HCMV-specific IgG activation of (E) FcγRIA and (F) FcγRIIA within paired maternal and cord blood sera. (G-H) Spearman correlations between HCMV-specific IgG signaling through FcγR and ADCP. Black bars denote median. P values for Mann-Whitney *U* test or Wilcoxon signed-rank test. * P < 0.05, ** P < 0.01, *** P < 0.001.

## Discussion

Despite decades of research, the immune correlates of protection against placental HCMV transmission have remained elusive and there are no licensed therapeutics or vaccines to prevent congenital HCMV infection (12). Using a case-control cohort of cord blood donor HCMV transmitting (n=41) and non-transmitting (n=40) mother-infant dyads, we identified Fc-mediated effector functions and ADCP as novel potential correlates of protection against congenital HCMV transmission. This finding represents a critical step forward towards the goal of preventing cCMV infection worldwide.

Consistent with our recent study on a subset of this cohort, HCMV-specific IgG transfer across the placenta was modestly decreased in cCMV infection and negatively correlated with maternal hypergammaglobulinemia (44). Despite lower transfer ratios, cord blood from cCMV-infected infants had high levels of HCMV-specific IgG, indicating that reduced transfer of HCMV-specific IgG across the placenta into the fetal circulation is not a risk factor for congenital infection. Since HCMV:IgG immune complex binding to FcRn has been proposed as a mechanism whereby HCMV can bypass the placental barrier (45, 52), we also examined whether HCMV-specific IgG binding to FcRn differed between transmitting and non-transmitting dyads. Normalized pentamer- and gHgLgO-IgG binding to FcRn was slightly higher in transmitting dyads (**Supplementary Figure 6A**), yet there were no differences within maternal-cord blood pairs to suggest enhanced placental transfer of anti-HCMV IgG via FcRn in the setting of congenital HCMV transmission.

Our finding that high-avidity HCMV-specific IgG was enriched in the cord blood of uninfected infants further confirms our previous work and indicates that high-avidity HCMV-specific IgG plays an important role in preventing congenital HCMV infection (44, 53). It is possible that the relative enrichment of low- avidity HCMV-specific IgG in the cord blood of infected infants is due to preferential transfer of low- avidity IgG across the placenta, which has been proposed as a mechanism of antibody-dependent enhancement (45, 52). However, we speculate that it is more likely an indication of recent maternal primary infection or reinfection leading to an abundance of low avidity antibodies present earlier in pregnancy that were then transferred to the fetus in the setting of congenital HCMV transmission. Our regression analyses and PCA of maternal antibody responses provide additional evidence that high-avidity HCMV-specific IgG is associated with protection against congenital infection (20, 23).

HCMV-specific IgG binding and neutralization have been correlated with protection against congenital transmission in some studies (21, 24), yet we unexpectedly found that HCMV glycoprotein-specific IgG binding and neutralizing titers were significantly higher in transmitting versus non-transmitting women. These seemingly discordant results are consistent with a recent study that observed higher gB- and pentamer-specific IgG levels in transmitting versus non-transmitting women with nonprimary infection (25). Dorfman et al. observed that IgG binding against putatively protective epitopes on pentamer were associated with elevated risk of congenital infection, which aligns with our finding that pentamer-specific IgG binding was associated with increased transmission risk. It has been proposed that elevated HCMV-specific IgG levels in transmitters are due to “boosting” from HCMV reactivation or reinfection (25). Nevertheless, we did not observe an association between maternal viral burden and HCMV-specific IgG levels or neutralizing titers, as the frequency of detection and levels of HCMV DNAemia were comparable in transmitting and non-transmitting mothers at the delivery timepoint (**Table 1**) . Moreover, HCMV-specific IgG binding and neutralizing titers remained significantly elevated in transmitters when excluding HCMV IgM positive mothers as a surrogate biomarker for recent primary infection or reinfection. Though pentamer IgG binding was associated with increased transmission risk, antibody-dependent enhancement of infection is unlikely given the compelling epidemiological and experimental evidence that CMV-specific IgG protects against placental transmission in human and animal studies (2, 54–56). Instead, we infer that boosted IgG levels and *in vitro* neutralizing titers in transmitting mothers are likely due to active HCMV infection following primary acquisition, reactivation, or reinfection during pregnancy.

Intriguingly, we found that ADCP, a non-neutralizing antibody effector function, was higher in non-transmitting compared to transmitting mother-infant dyads and that maternal ADCP responses were associated with decreased risk of congenital HCMV infection. Moreover, sera from non-transmitting dyads demonstrated increased HCMV-specific IgG activation of FcγRI and FcγRIIA, the FcγRs on monocytes, macrophages, and dendritic cells that mediate ADCP (49, 57, 58). Mechanistically, these findings indicate that enhanced FcγRI and FcγRIIA engagement by HCMV-specific IgG mediates non- neutralizing antibody responses that protect against placental HCMV transmission. Given that HCMV infection primarily spreads cell-to-cell *in vivo*, it is not altogether surprising that we found that non- neutralizing antibody responses were associated with protection against *in utero* HCMV transmission (59). ADCP can encompass phagocytosis of IgG:virus immune complexes as well as phagocytosis of infected cells presenting viral antigens and therefore can defend against both cell-associated and cell- free virus (57, 60), making it a logical protective immune response against HCMV infection. Notably, FcγRI and FcγRIIA are highly expressed on maternal- and fetal-derived monocytes/macrophages at the maternal-fetal interface, as revealed by several recent single-cell studies (61–63). Thus, it is intriguing to consider whether these innate immune cells employ Fc-mediated effector functions to protect against placental HCMV infection directly (39). Future studies should explore whether other Fc-mediated effector functions in addition to ADCP protect against congenital HCMV transmission either systemically or at the maternal-fetal interface.

Our findings that ADCP and FcγR engagement are correlated with protection against placental HCMV transmission have important implications for HCMV vaccine development beyond the context of congenital infection. The most successful HCMV vaccine to-date was a gB subunit vaccine with an MF59 adjuvant that achieved ∼50% efficacy in HCMV seronegative women and solid organ transplant recipients, yet the functional antibody responses mediating this protection have remained elusive (35, 37, 38, 64). Several studies have demonstrated that neutralizing antibodies were poorly elicited by the gB/MF59 vaccine, leading researchers to hypothesize that non-neutralizing antibody responses mediated vaccine efficacy (34, 36, 38). Though non-neutralizing ADCC responses were poorly stimulated in gB/MF59 vaccinees, ADCP against HCMV virions and gB-coated beads were robustly induced (34, 36, 38). While ADCP was not identified as an immune correlate of protection against HCMV infection in our recent study of adolescent and postpartum gB/MF59 vaccinees, there was a trend towards ADCP being significantly associated with protection against virus acquisition (β = -0.420, p = 0.120) (36). Notably, IgG binding to cell-associated gB, which we identified as an immune correlate of protection for gB/MF59 vaccinees, was significantly correlated with HCMV-specific ADCP and FcγRI/FcγRIIA activation in our study (**Supplementary Figure 4**). Cumulatively, these results suggest that non-neutralizing IgG responses against gB in a native conformation on the surface of a virion or cell may protect against HCMV in both natural infection and vaccination. Taken together, our data indicates that non-neutralizing Fc effector functions, whether mediated by gB-specific IgG or other IgG specificities, should be investigated as putative correlates of protection in future HCMV vaccine trials including the ongoing Moderna gB/pentamer mRNA phase 3 clinical trial (ClinicalTrials.gov identifier NCT05085366). Notably, antibodies eliciting Fc effector functions can be polyfunctional, mediating both neutralizing and non-neutralizing functions (65–68). Therefore, eliciting neutralizing and non-neutralizing antibodies should not be viewed as mutually exclusive in future HCMV vaccine trials.

Different biophysical and biochemical properties of IgG including subclass and glycosylation modify FcγR binding affinity and subsequent Fc effector functions (69). For instance, in HIV vaccine trials, HIV- specific IgG3 binding to activating FcγRs has been linked with enhanced ADCP and ADCC and correlated with vaccine efficacy (65, 66). While in severe COVID-19, afucosylation of the Fc region of anti-SARS-CoV-2 IgG antibodies enhances binding to FcγRIIIA, which significantly modifies downstream pro-inflammatory cytokine signaling (70). Thus, future studies of immune correlates and mechanisms of protection against HCMV infection and vaccine efficacy should also investigate how differences in IgG subclass or Fc glycosylation may be modulating Fc-mediated antibody responses.

Our study has several limitations. Due to the cross-sectional and retrospective nature of this cord blood bank donor mother-infant cohort, we were unable to definitively identify maternal primary versus nonprimary HCMV infections and the timing of maternal infection. Placental biospecimens and maternal PBMCs were not collected by the CCBB, so we could not investigate placental infection nor identify maternal cellular immune correlates of protection, even though CMV-specific T cell responses have been associated with protection against placental transmission (23, 33, 71). Since long-term clinical outcomes were unavailable, we could not assess whether antibody responses correlated with protection against long-term disease sequelae, though all infants were born without symptomatic disease at birth. Statistical power was limited by small sample size (n = 41 HCMV transmitting dyads), yet this represents one of the largest U.S.-based cohorts used to assess congenital HCMV immune correlates to-date and one of the few studies that does not have the confounder of maternal HIV co-infection (21, 23–26, 72). Our study focused on ADCP and did not measure other Fc effector functions such as ADCC, antibody-dependent neutrophil phagocytosis (ADNP), and antibody-dependent complement deposition (ADCD) due to limited sera sample volumes. However, future studies should investigate whether these Fc effector functions also mediate protection against cCMV transmission.

Development of an efficacious HCMV vaccine has been considered a “top tier priority” by the U.S. National Academy of Medicine for over 20 years, but identifying immune correlates of protection against maternal or congenital HCMV infection has proved challenging, hindering vaccine progress to-date (73, 74). Though gB- and pentamer-specific IgG binding titers and neutralizing antibodies have been the main focus of HCMV vaccine development (10, 33), these responses were not associated with protection against congenital transmission in our cohort. Instead, our study suggests that eliciting HCMV-specific IgG that engages FcγRI/FcγRIIA and mediates non-neutralizing Fc effector functions such as ADCP represents an important immune response in the prevention of congenital HCMV transmission and a promising new approach for HCMV vaccinology. These findings will guide the ongoing development of active and passive maternal immunization strategies with the goal of preventing cCMV transmission in seropositive and seronegative pregnant women worldwide.

## Methods

### Study population

We analyzed maternal and cord blood sera from 81 mother-infant dyads recruited from 2008-2017 as donors to the Carolinas Cord Blood Bank (CCBB). The CCBB collects maternal and cord blood biospecimens at delivery and mother-infant dyads were identified from over 29,000 CCBB donor records (**Supplementary Figure 1**). Maternal donors underwent infectious diseases screening for HCMV, hepatitis B virus, syphilis, hepatitis C virus, HIV-1/2, HTLV I and II, Chagas Disease, and West Nile virus. All mothers in our study were HCMV IgG seropositive and negative for other infectious diseases. Cord blood plasma was screened by the CCBB for HCMV infection with a Real-Time PCR COBAS AmpliPrep/TaqMan nucleic acid test (WHO HCMV reference standard, limit of quantification 147 IU/mL (75)), and all cord blood units positive for HCMV underwent a second confirmatory PCR test.

Cases of congenital HCMV infection (cCMV) were defined as mother-infant dyads with cord blood that screened positive for HCMV viremia at birth per PCR testing. “HCMV transmitting” cases with cCMV infection (n=41) were matched to a target of 1 “HCMV non-transmitting” seropositive mother (n=40) with no HCMV viremia detected in the cord blood (**Supplementary Figure 1**). Maternal HCMV IgG seropositivity and avidity were confirmed by an in-house whole virion HCMV ELISA and HCMV IgM seropositivity was determined using a clinical diagnostic ELISA (Bio-Rad). Maternal sera was screened for HCMV DNAemia via quantitative PCR (qPCR). For qPCR, 150-200ul of maternal sera was ultracentrifuged then DNA was extracted using the DNA QIAamp Kit (Qiagen) and tested in duplicate using SybrSelect and 300nM of forward and reverse primers designed to amplify the HCMV immediate early-1 (IE1) gene (24). HCMV viral loads were then interpolated from an IE1 plasmid standard curve. Maternal sera was also screened for hypergammaglobulinemia (total IgG concentration >15,000 mg/dL) via ELISA as previously described (44).

### Cell culture and HCMV virus growth

Human retinal pigment epithelial cells (ARPEs), human foreskin fibroblasts (HFFs), human embryonic kidney (HEK)-293T cells, and human monocytes (THP-1s) were acquired from ATCC and cultured according to ATCC protocols. For differentiation, THP-1 cells were cultured in R10 media with 100nM PMA and incubated for 48 hours. HCMV strains TB40/E, AD169r, and Toledo were propagated as previously described (76). HFF or ARPE cells were infected (MOI = 0.01) and incubated for ∼14 days until 90-95% of cells showed cytopathic effect. Cells and supernatant were harvested, filtered (0.45-μm), and then concentrated on a 20% sucrose cushion (20,000 rpm).

### Whole virion HCMV IgG binding and avidity

384-well ELISA plates were coated with 33 PFU/well of TB40/E, 2700 PFU/well of AD169r, or 1000 PFU/well of Toledo virus diluted in 0.1 M sodium bicarbonate buffer then incubated overnight before blocking. Maternal and cord blood sera serial dilutions were tested starting at 1:30, plated in duplicate, then incubated before adding horseradish peroxidase (HRP)-conjugated goat anti-human IgG Fc (Jackson ImmunoResearch). Plates were developed with tetramethylbenzidine (TMB) and peroxidase substrate (KPL) then optical density (OD) at 450nm was measured via SpectroMax. HCMV-specific IgG concentrations were interpolated from the linear range of a 5-parameter HCMV-hyperimmuneglobulin (Cytogam) standard curve. For IgG avidity, duplicate wells were treated with 7M urea or 1X PBS between the primary and secondary incubation steps and relative avidity index (RAI) was calculated as (OD with urea)/(OD with PBS)x100%. Placental HCMV-specific IgG transfer was calculated as (cord blood IgG)/(maternal IgG)x100%. Duplicates with coefficients of variance (CVs) >20% were repeated.

### HCMV glycoprotein-specific IgG binding and avidity

A binding antibody multiplex assay (BAMA) was used to quantify HCMV glycoprotein-specific IgG binding and avidity (76, 77). HCMV gB ectodomain, pentamer complex, gH/gL/gO and gH/gL antigens were covalently coupled to intrinsically fluorescent beads (Bio-Rad). Maternal and cord blood sera were diluted 1:500 in assay diluent, plated in duplicate, then co-incubated with antigen-coupled beads. Antigen-specific IgG binding was detected with mouse anti-human IgG-PE (Southern Biotech) and mean fluorescent intensity (MFI) was acquired on a Bio- Plex 200 (Luminex). To quantify avidity, duplicate wells were incubated with sodium citric acid (pH = 4.0) or 1X PBS (pH = 7.4) between the primary and secondary incubations and RAI was calculated as (MFI with sodium citric acid)/(MFI with PBS)x100%. A serial dilution of HCMV-hyperimmuneglobulin (Cytogam) was included as a positive control and the cut-off for positivity was determined by calculating the mean MFI of seronegative sera binding plus 3 standard deviations. Blank beads and wells were included to account for background signal. Duplicates with CVs >25% were repeated.

### Fc receptor (FcR) binding by HCMV glycoprotein-specific IgG

FcR binding by HCMV glycoprotein- specific IgG was measured using a modified BAMA (36, 78). Purified human FcRn, FcR1a, FcR2a (clones H131 and R131), FcR2b, and FcR3a (clones V158 and F158) were produced by the DHVI Protein Production Facility and biotinylated in-house. Maternal and cord blood sera were diluted 1:500 in assay diluent before incubation with HCMV glycoprotein-coated beads, as above. Next, biotinylated human FcRs were complexed with streptavidin-PE (BD Biosciences) then co-incubated with antibody- bound beads after washing. For FcRn, the assay buffer pH was adjusted to 6.5 to facilitate FcRn-IgG binding. MFI was acquired on a Bio-Plex 200 and duplicates with CVs >25% were repeated.

### Cell-associated HCMV glycoprotein B (gB) IgG binding

A gB-transfected cell binding assay was used to measure IgG binding to cell-associated gB (36). HEK-293T cells were co-transfected with DNA plasmids expressing green fluorescent protein (GFP) and full-length gB (Sino Biological) using the Effectene Transfection Kit (Qiagen). After incubation at 37°C for 48 hours, 200,000 live cells/well were plated into 96-well U-bottom plates then centrifuged and washed before a 5 minute incubation in Human TruStain Fc Block (1:1000 dilution; BioLegend). In duplicate, cells were then co-incubated with maternal and cord blood sera diluted 1:500 or with controls for 2 hours at 37°C. A serial dilution of HCMV-hyperimmuneglobulin (Cytogam) and a gB-specific monoclonal antibody (TRL-345) were included as positive controls and seronegative sera samples were included as negative controls. Following incubation, cells were stained with Live/Dead Near-IR (1:1000; Invitrogen) then washed and stained with PE-conjugated goat anti-human IgG-Fc (1:200; Southern Biotech). Lastly, cells were washed and fixed with 10% Formalin. Events were acquired on an LSRII flow cytometer, and the percentage of PE-positive cells was calculated from the live, GFP-positive cell parent population using FlowJo (36). Unstained cells and single color-stained cells were included for setting gates and compensation. The cut-off for positivity was the mean signal of HCMV seronegative samples plus 3 standard deviations. Duplicates with CVs >50% were repeated.

### Neutralization

HCMV neutralization was measured by high-throughput fluorescence bioimaging (36). Epithelial cells (ARPEs), fibroblasts (HFFs) or differentiated monocytes/macrophages (THP-1s) were plated in 384-well clear, flat-bottom plates and then incubated at 37°C overnight. To quantify neutralization, maternal and cord blood sera samples were diluted 1:10 followed by an 8-point serial dilution and then co-incubated with HCMV strains AD169r (MOI = 2) or Toledo (MOI = 1) for 2 hours at 37°C to allow immune complex formation. Virus-only wells and an 8-point HCMV-hyperimmuneglobulin (Cytogam) serial dilution were included as positive controls while seronegative samples and no virus wells were included as negative controls. After addition of the virus:sera mixture, cells were incubated at 37°C for 24-48 hours (depending on optimized conditions for each virus strain and cell type) then fixed in 10% Formalin. To quantify HCMV infection, plates were stained with mouse anti-HCMV immediate-early 1 (IE1) gene (1:500; MAB810 Millipore) followed by goat anti-mouse IgG-AF488 (1:500; Millipore) and cell nuclei were stained with DAPI (1:10,000; Thermo Fisher). After staining, plates were visualized with a Cellomics fluorescent plate reader (**Supplementary Figure 8A**) to enumerate total cell count, infected cell count, and percent infected cells in each well. Following image acquisition, neutralization titers corresponding to the dilution that resulting in a 50% reduction in percent infected cells (ID50) were calculated using interpolation in GraphPad Prism (**Supplementary Figure 8B**). Samples containing wells with low cell counts and with duplicates with CVs >50% were repeated.

### Whole HCMV virion antibody-dependent cellular phagocytosis (ADCP)

HCMV strains TB40/E, AD169r, and Toledo were conjugated to fluorochrome AF647 (Invitrogen) to measure ADCP (36). In brief, 2.0×10^6^ PFU of TB40/E, 1.0×10^7^ PFU of AD169r, or 1.0×10^7^ PFU of Toledo virions were buffer-exchanged with 1X PBS using a 100,000-kDa Amicon filter (Millipore) and conjugated to 10µg of AF647 N-hydroxysuccinimide ester reconstituted in DMSO during a 1-hour incubation with constant agitation. The conjugation reaction was quenched with 80ul of 1M tris-HCl (pH = 8.0) and fluorescently-labeled virus was diluted 25X in wash buffer. A serial dilution of HCMV-hyperimmuneglobulin (Cytogam) was included as a positive control while seronegative sera samples and an anti-RSV monoclonal antibody (Synagis) were included as negative controls. In a 96-well plate, fluorescently-labeled virus was co- incubated with maternal sera, cord blood sera (1:10) or controls at 37°C for 2 hours to allow immune complex formation before adding 50,000 THP-1 cells per well. Plates were then centrifuged (1200g) at 4°C for 1 hour in a spinoculation step before a 1-hour incubation at 37°C to allow for phagocytosis. Next, cells were transferred to a 96-well U-bottom plate, washed and fixed with 1% Formalin. Events were acquired on an LSRII flow cytometer, and the percentage of AF647-positive cells was calculated from the live THP-1 monocyte cell parent population using FlowJo (gating strategy in **Supplementary Figure 9C-D**). Unstained cells and single color-stained cells were included as controls for setting gates and compensation. The cut-off for positivity was the mean signal of HCMV seronegative samples plus 3 standard deviations and duplicates with CVs >50% were repeated.

### Fcγ receptor signaling assay

HCMV-specific IgG signaling through FcγRs was measured using a previously validated approach (50). Briefly, we used mouse BW thymoma cells stably expressing chimeric FcR-CD3ζ, which contains an extracellular human FcR and intracellular CD3ζ signaling domain, to quantify anti-viral IgG activation of host FcγRs. To confirm FcγR expression, 0.5×10^6^ BW cells were added to each well in a 96-well V-bottom plate. The non-transfected parental BW cell line and BW cell lines expressing either human CD64 (FcR1a) or CD32a (FcR2a) were stained for surface expression of human FcγRs with 5µl of anti-human CD64-PE (clone 10.1, eBioscience), 5µl anti-human CD32 (clone 6C4, eBioscience), and 5µl anti-human Ig-PE isotype control (clone P3.6.2.8.1, eBioscience) then cells were fixed with 2% paraformaldehyde. Events were acquired on Fortessa LSR flow cytometer and analyzed using FlowJo v10.7.2.

To quantify FcγR activation, 96-well plates were coated with 20,000 PFU/well of HCMV strain AD169r and incubated at 4°C overnight. After coating, plates were washed with assay buffer (1X PBS + 1% FBS) then blocked with buffer at room temperature for 1 hour. After blocking, HCMV-coated plates were co-incubated with maternal or cord blood sera diluted 1:10 in BW cell media at 37°C for 1 hour. HCMV-hyperimmuneglobulin (Cytogam), seronegative and no antibody conditions were included as positive and negative controls. Following immune complex formation, plates were washed with BW media before adding 100,000 FcR1a-expressing or FcR2a-expressing BW cells per well. In separate wells, the parental (non-transfected) BW cells were added as a negative control. Cells were incubated at 37°C with immune complex coated plates for 20 hours before being transferred to V-bottom plates and pelleted (1200 rpm). Cell supernatants were harvested and mouse IL-2 (mIL-2) levels in culture supernatants were measured using ELISA. For the mIL-2 ELISA, 384-well plates were coated with 3µg/mL of purified rat anti-mouse IL-2 (BD biosciences) and incubated at 4°C overnight before blocking. A serial dilution of purified mIL2 and BW cell culture supernatants were added in duplicate and incubated for 1 hour at room temperature. After primary incubation, plates were incubated with rat anti-mIL2 conjugated to biotin (BD biosciences; 1:2000) followed by streptavidin-HRP (1:8000) for 1 hour and 30 minutes respectively. Plates were developed with TMB/KPL then OD at 450nm was measured via SpectroMax. Mouse IL-2 concentrations were interpolated from a 5-parameter mIL-2 standard curve using GraphPad Prism. Duplicates with CVs >30% were repeated.

### Statistics

All primary raw and analyzed data underwent independent data quality control (QC) by a second lab member prior to statistical analysis and inclusion in the study. For all analyses, interpolated antibody or cytokine concentrations, MFI values and neutralization titers were log-transformed to normalize the data distribution. Antibody responses below the limit of detection were set equal to the limit of detection for statistical analyses. To assess differences between HCMV transmitting and non- transmitting mother-infant dyads, immune variables were compared between groups using Mann- Whitney U/Wilcoxon rank-sum and within dyads using Wilcoxon signed rank tests. Statistical significance was defined a priori as p < 0.05 with a two-tailed test and a Benjamini-Hochberg correction for multiple comparisons. For the primary immune correlate analysis, 13 predefined maternal humoral immune variables were included for univariate logistic regression analysis. Additional sensitivity analyses included multivariable models adjusted for 1) maternal hypergammaglobulinemia and 2) maternal HCMV-specific IgM status. Using the 13 predefined immune variables from our primary immune correlate analysis, LASSO regression analysis was performed using the caret R package and a k-fold cross-validation framework. For the LASSO regression, the cohort was randomly split into two independent datasets, which were used for training the LASSO model and testing the predictive performance of the model respectively. All statistical analyses were completed in R v4.1.1 and GraphPad Prism v9.1. The principal components analysis (PCA) plots were rendered using ggplot2 and correlation matrices were plotted using the *corrplot* package in R. All other figures were generated using GraphPad Prism or created in Biorender.com.

### Study Approval

Approval was obtained from Duke’s Institutional Review Board (Pro00089256) to use de-identified clinical data and biospecimens provided by the CCBB. No patients were prospectively recruited for this study and all samples were acquired retrospectively from the CCBB biorepository from donors who had previously provided written consent for banked biospecimens to be used for research.

## Author contributions

ECS, JAJ, JHH, KMW, and SRP designed the research study. ECS, IGM, SJB, MJH and HW conducted the experiments and acquired the data. ECS, IGM and SJB completed the primary data analysis. CEW and ECS completed the statistical analyses with oversight from KMW. SRP and KMW acquired funding for the study. JK provided the biospecimens and clinical data for the study. ECS wrote the primary draft of the manuscript. ECS, IGM, JAJ, CEW, SJB, MJH, HW, JHH, JK, GGF, KMW and SRP all contributed to writing and editing the manuscript.

## Data Availability

All data produced in the present study are available upon reasonable request to the authors.

## Acknowledgements

Thank you to the CCBB donors and CCBB staff including Jose Hernandez, Ann Kaestner, and Korrynn Vincent who were instrumental in acquiring the biospecimens and donor clinical information for this study. We also would like to thank Drs. Phillip Kolb and Hartmut Hengel who generously provided the FcR-transfected BW cell lines. This project was supported by NIH NCI 1R21CA242439-01 “Immune Correlates and Mechanisms of Perinatal Cytomegalovirus Infection and Later Life ALL Development” to KMW and SRP and NIH NIAID 1R21-AI147992 “Humoral immune correlates of protection against congenital CMV and HSV transmission in HIV-infected women” to SRP. Additional support was provided by Duke University School of Medicine through Translating Duke Health’s Children’s Health and Discovery Initiative and the Medearis CMV Scholars Program to SRP. The funders had no role in study design, data collection and analysis, decision to publish, or preparation of the manuscript.

