## Supplementary Figures and Tables for "Maternal Fc-mediated non-neutralizing antibody responses correlate with protection against congenital human cytomegalovirus infection"

**Supplementary Figure 1.** Identification of HCMV transmitting and non-transmitting mother-infant dyads from the Carolinas Cord Blood Bank (CCBB) biorepository.

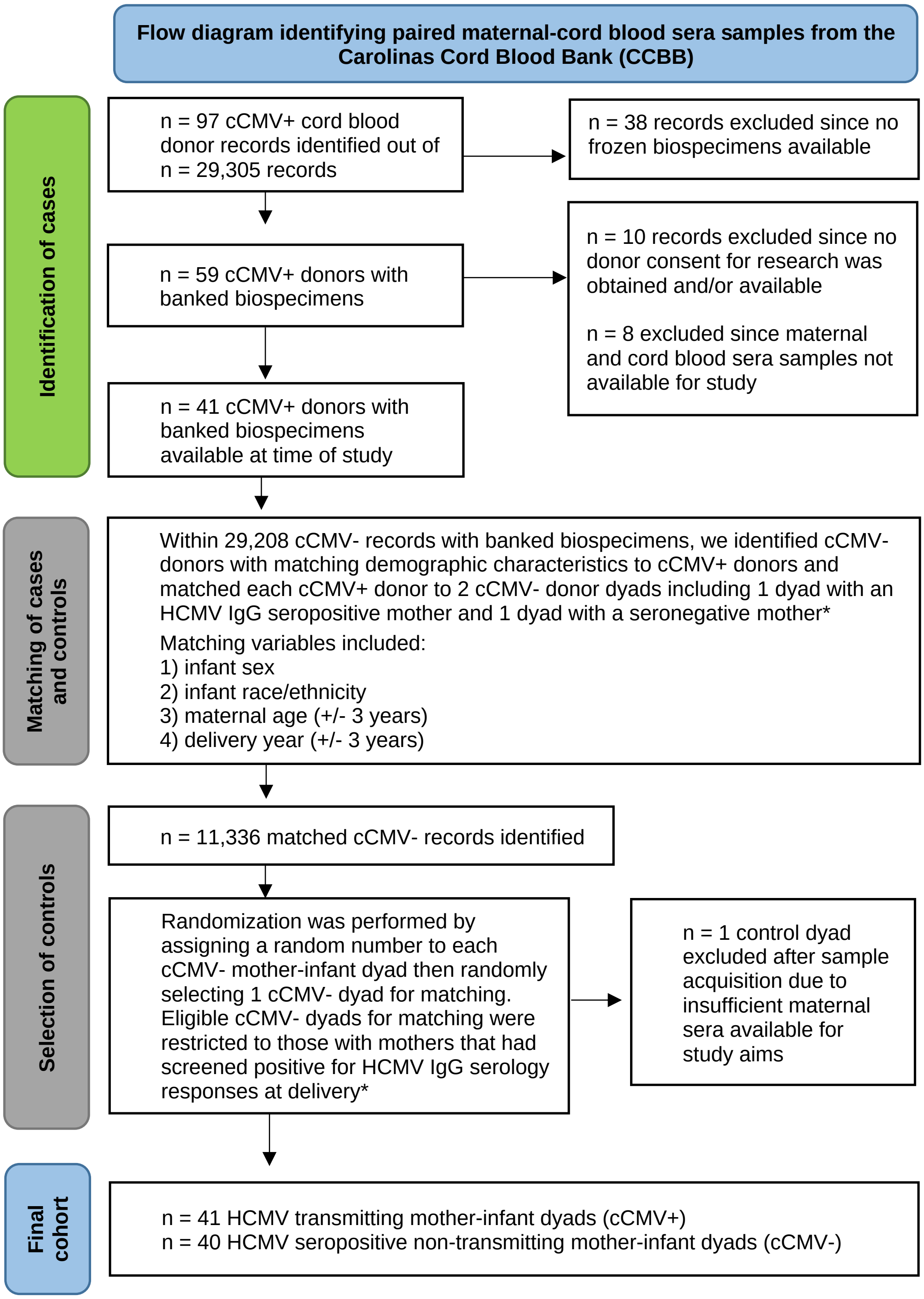


cCMV = congenital HCMV infection
cCMV+ = positive HCMV PCR cord blood screening at birth
cCMV- = negative HCMV PCR cord blood screening at birth

*Initial HCMV serology screening performed at time of donation by the American Red Cross in Charlotte, N.C.

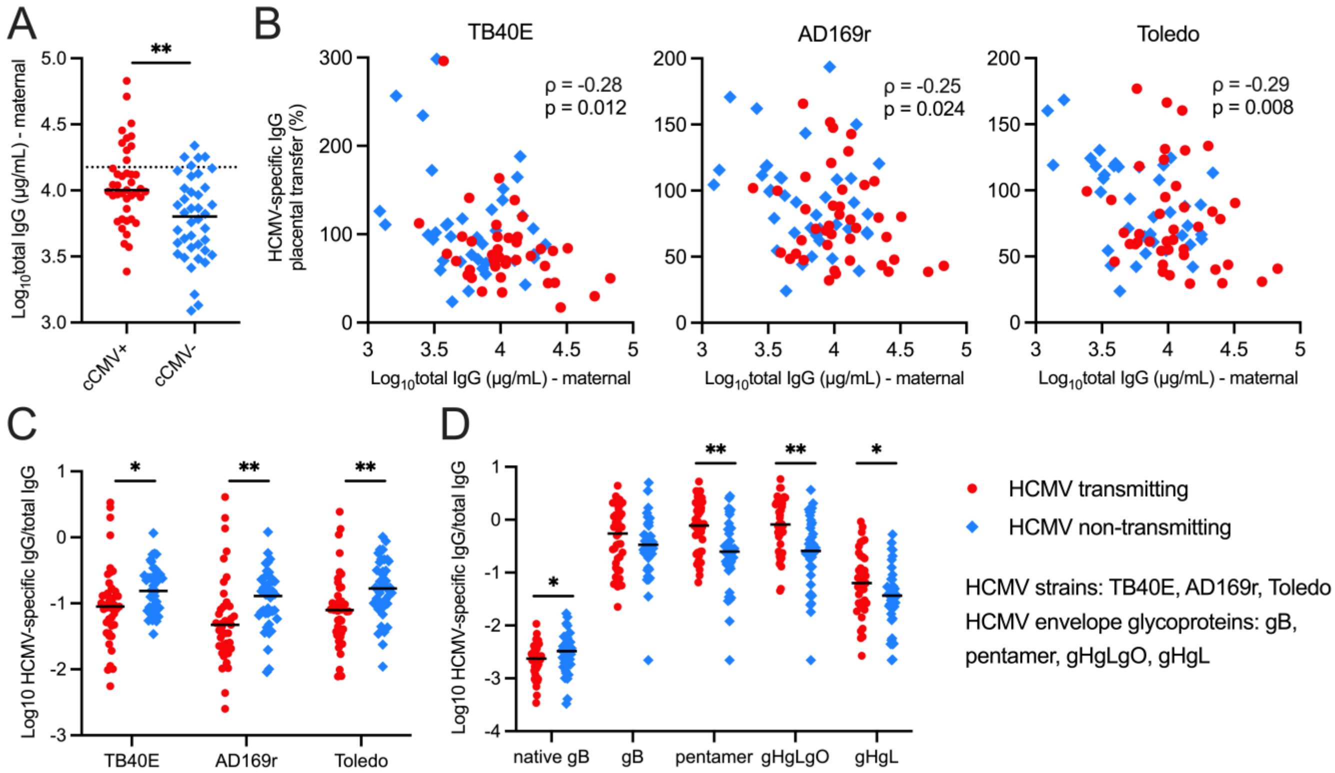


**Supplementary Figure 2. Maternal hypergammaglobulinemia in HCMV transmitting dyads correlates with decreased placental HCMV-specific IgG transfer efficiency.** Total IgG levels and HCMV-specific IgG binding against HCMV strains TB40E, AD169r, and Toledo were measured using ELISA. Transplacental transfer efficiency of HCMV-specific IgG was calculated as (cord blood HCMV-specific IgG concentration)/(maternal HCMV-specific IgG concentration)x100%. Cell-associated gB IgG binding was quantified using a flow-based assay and HCMV glycoprotein-specific IgG binding magnitude was measured using a Luminex-based binding antibody multiplex assay. Antibody responses in maternal and cord blood sera were compared between HCMV transmitting (red circles, n = 41) and non-transmitting (blue squares, n = 40) mother-infant dyads. (A) Maternal sera total IgG levels in transmitting versus non-transmitting dyads. Dotted line indicates cut-off for hypergammaglobulinemia (total IgG concentration >15,000 mg/dL). (B) Spearman correlations between HCMV-specific IgG placental transfer efficiencies and total maternal IgG levels in transmitting and non-transmitting dyads. (C-D) Maternal sera HCMV-specific IgG binding normalized to maternal total IgG levels in transmitting versus non-transmitting dyads. Black bars denote median. P values reported for Mann-Whitney *U* test or Spearman rank correlation. * P < 0.05, ** P < 0.01, *** P < 0.001.

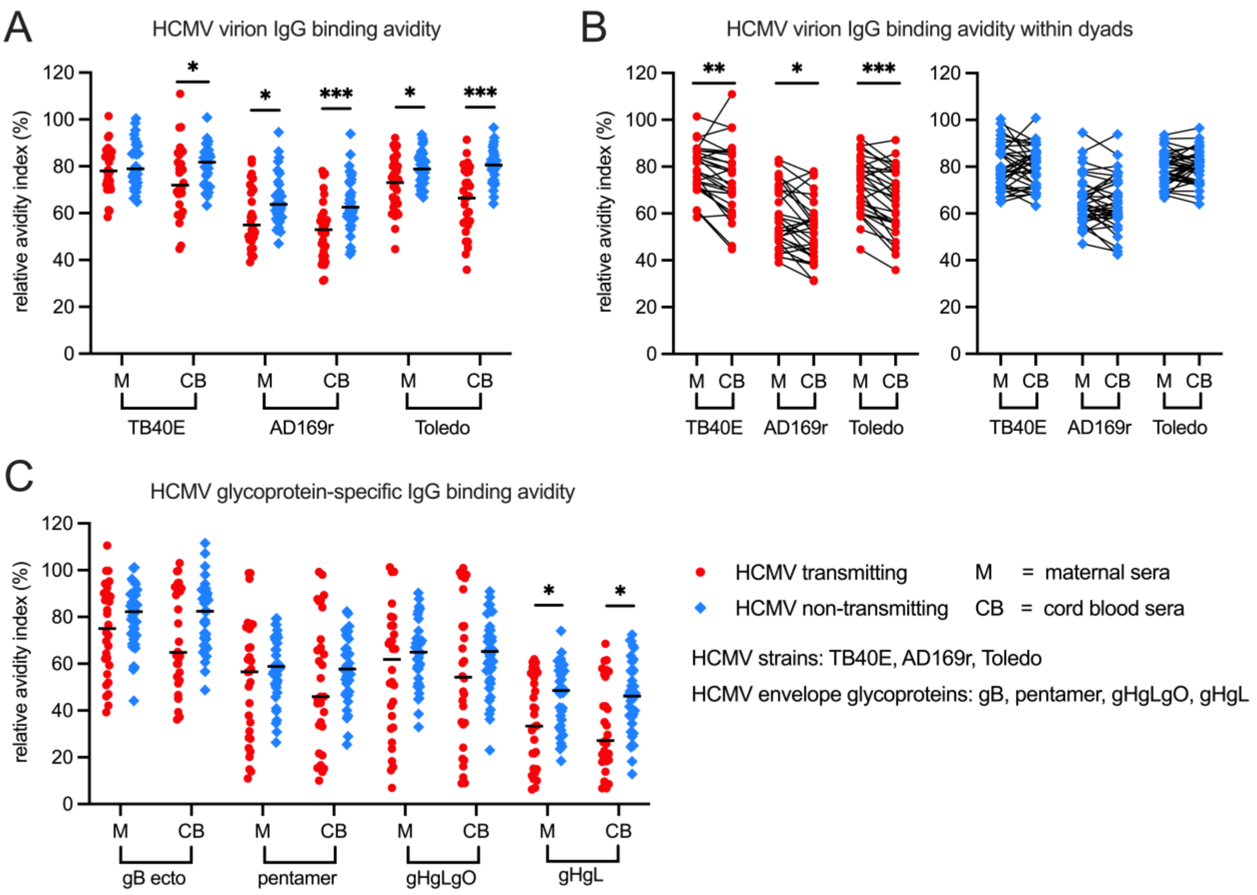


**Supplementary Figure 3. Higher HCMV-specific IgG avidity in HCMV transmitting versus non-transmitting dyads persists when excluding mothers with detectable HCMV-specific IgM responses.** HCMV-specific IgG binding avidity against HCMV strains TB40E, AD169r, and Toledo were measured using whole virion ELISA with an additional dissociation step using urea and relative avidity index (RAI) was calculated as (OD with urea/OD without urea)X100%. HCMV glycoprotein-specific IgG binding avidity was measured using a binding antibody multiplex assay with an additional dissociation step with sodium citrate buffer (pH = 4) and RAI was calculated as (MFI with sodium citrate/MFI with 1X PBS)X100%. In a sensitivity analysis excluding dyads where mothers screened positive for HCMV-specific IgM responses as a surrogate for recent primary infection or reactivation, IgG binding avidity in maternal (M) and cord blood (CB) sera was compared between and within a subset of transmitting (red circles, n = 30) and non-transmitting (blue squares, n = 38) mother-infant dyads. (A) Whole virion HCMV-specific IgG binding avidities in transmitting versus non-transmitting dyads. (B) Whole virion HCMV-specific IgG binding avidities in paired maternal and cord blood sera. (C) HCMV glycoprotein-specific IgG binding avidities in transmitting versus non-transmitting dyads. gB ecto = gB ectodomain. Black bars denote median. FDR-corrected P values for Mann-Whitney *U* test or Wilcoxon signed-rank test. * P < 0.05, ** P < 0.01, *** P < 0.001.

**Supplementary Table 1. Univariate logistic regression analysis of maternal humoral immune correlates of placental HCMV transmission adjusted for potential confounders.**

| | Univariate adjusted for maternal total IgG | | Univariate corrected for maternal HCMV-specific IgM status | |
|---|---|---|---|---|
| **Antibody response** | **Beta coefficient**[b] | ***P* value**[c] | **Beta coefficient**[b] | ***P* value**[c] |
| **IgG binding to cell-associated gB (%)** | **0.187** | **0.007** | **0.187** | **0.007** |
| **gB ectodomain IgG binding (Log MFI)** | **0.653** | **0.006** | **0.653** | **0.006** |
| **pentamer IgG binding (Log MFI)** | **1.712** | **<0.0001** | **1.712** | **<0.0001** |
| **gHgLgO IgG binding (Log MFI)** | **1.431** | **<0.0001** | **1.431** | **<0.0001** |
| **gB IgG avidity (%)** | **-0.027** | **0.049** | **-0.027** | **0.049** |
| **pentamer IgG avidity (%)** | -0.018 | 0.106 | -0.018 | 0.106 |
| **gHgLgO IgG avidity (%)** | -0.022 | 0.050 | -0.022 | 0.050 |
| **Fibroblast neutralization (Log ID50)**[a] | **0.536** | **0.042** | **0.536** | **0.042** |
| **Epithelial neutralization (Log ID50)**[a] | **1.328** | **0.002** | **1.328** | **0.002** |
| **Macrophage neutralization (Log ID50)**[a] | **1.533** | **0.000** | **1.533** | **0.000** |
| **Whole virion Toledo ADCP (%)** | **-0.044** | **0.046** | **-0.044** | **0.046** |
| **Whole virion TB40E ADCP (%)** | -0.023 | 0.254 | -0.023 | 0.254 |
| **Whole virion AD169r ADCP (%)** | -0.032 | 0.475 | -0.032 | 0.475 |

[a] Neutralization measured against HCMV strain AD169r

[b] Positive beta coefficients are associated with increased risk and negative beta coefficients are associated with decreased risk of placental HCMV transmission

[c] **Bold indicates statistical significance (*P* value <0.05)**

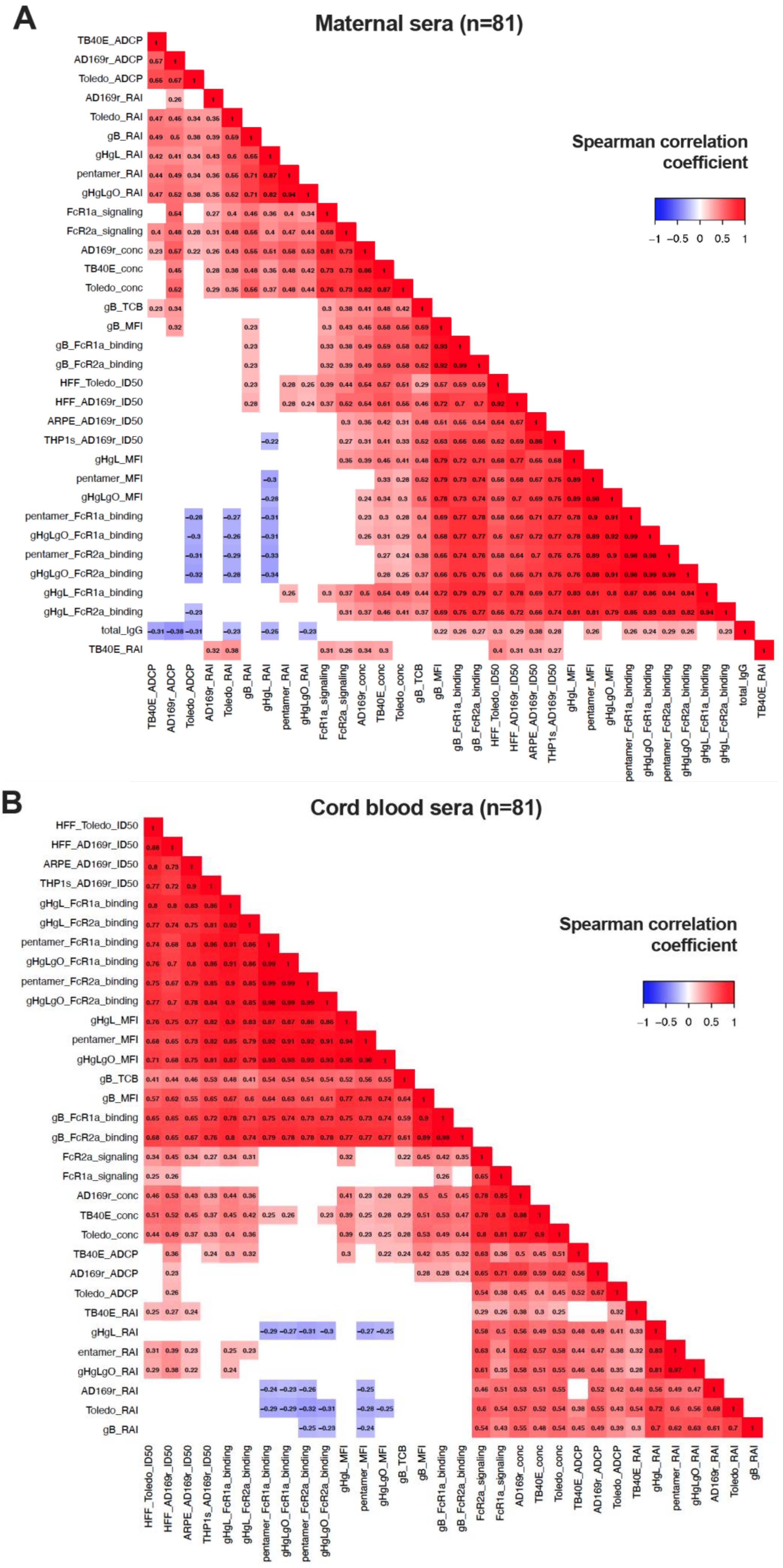


**Supplementary Figure 4. Correlation matrix of HCMV-specific antibody responses.** Hierarchical clustering was performed on Spearman correlation coefficients to group strongly correlated immune variables. Matrix of antibody responses showing Spearman correlation coefficients from -1.0 (blue) to +1.0 (red) in (A) maternal and (B) cord blood sera. Non-significant correlations (uncorrected P > 0.05) shown in white. gB_TCB = cell-associated gB transfected cell binding.

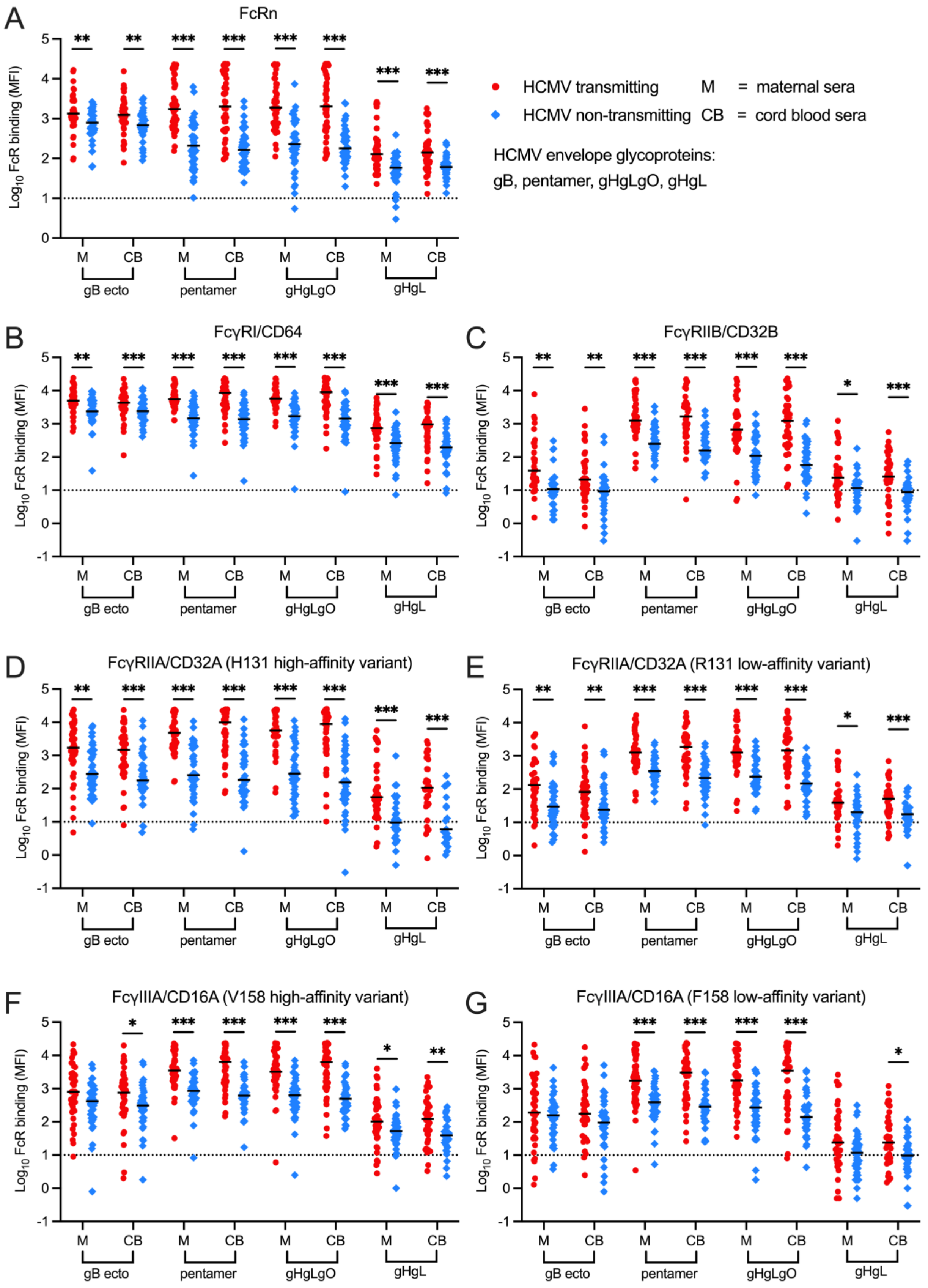


**Supplementary Figure 5. Non-normalized HCMV glycoprotein-specific IgG binding to Fc receptors in transmitting versus non-transmitting dyads.** HCMV glycoprotein-specific IgG binding to (A) neonatal Fc receptor (FcRn), (B) FcγRI/CD64, (C) FcγRIIB/CD32B, (D-E) FcγRIIA/CD32A high-affinity (H131) and low affinity (R131) variants, and (F-G) FcγRIIIA/CD16A high affinity (V158) and low affinity (F158) variants in maternal (M) and cord blood (CB) sera was compared between transmitting (red circles, n = 41) and non-transmitting (blue squares, n = 40) mother-infant dyads. Black bars denote median. Dotted line indicates lower limit of detection. FDR-corrected P values for Mann-Whitney *U* test. * P < 0.05, ** P < 0.01, *** P < 0.001.

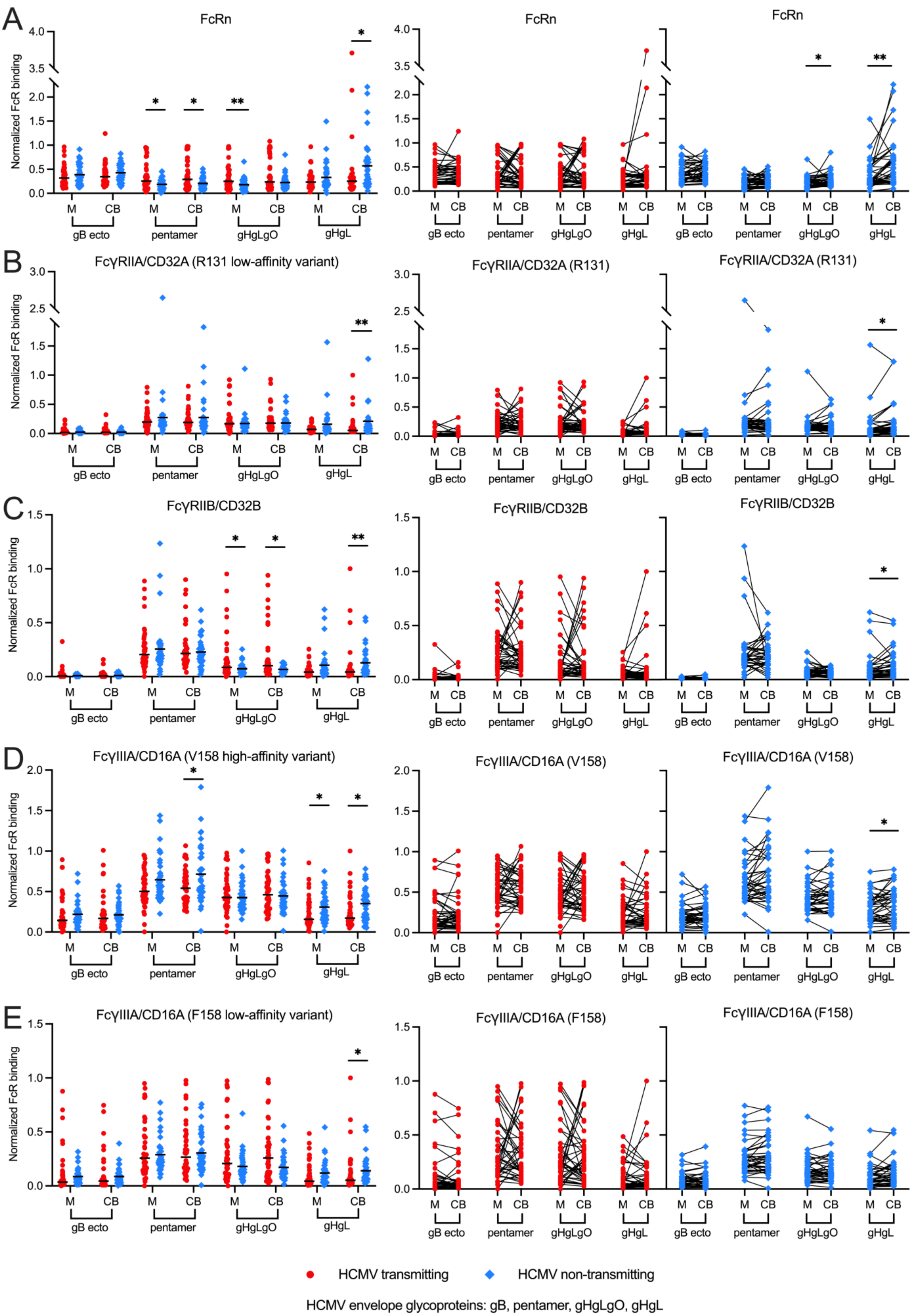
A
FcRn
Normalized FcR binding
M
CB
gB ecto
pentamer
gHgLgO
gHgL
B
FcγRIIA/CD32A (R131 low-affinity variant)
FcγRIIA/CD32A (R131)
C
FcγRIIB/CD32B
D
FcγIIIA/CD16A (V158 high-affinity variant)
FcγIIIA/CD16A (V158)
E
FcγIIIA/CD16A (F158 low-affinity variant)
FcγIIIA/CD16A (F158)
HCMV transmitting
HCMV non-transmitting
HCMV envelope glycoproteins: gB, pentamer, gHgLgO, gHgL

**Supplementary Figure 6 (see previous page). Normalized HCMV glycoprotein-specific IgG binding to Fc receptors in transmitting and non-transmitting dyads.** HCMV glycoprotein-specific IgG binding to (A) neonatal Fc receptor (FcRn), (B) FcγRI/CD64, (C) FcγRIIB/CD32B, (D-E) FcγRIIA/CD32A high-affinity (H131) and low affinity (R131) variants, and (F-G) FcγRIIIA/CD16A high affinity (V158) and low affinity (F158) variants in maternal (M) and cord blood (CB) sera was normalized to total sera IgG binding to gB-, pentamer-, gHgLgO- and gHgL-coated multiplex beads at baseline in binding antibody multiplex assay. Normalized IgG binding to FcRs was compared between and within transmitting (red circles, n = 41) and non-transmitting (blue squares, n = 40) mother-infant dyads. gB ecto = gB ectodomain. Black bars denote median. FDR-corrected P values for Mann-Whitney *U* test or Wilcoxon signed-rank test. * P < 0.05, ** P < 0.01, *** P < 0.001.

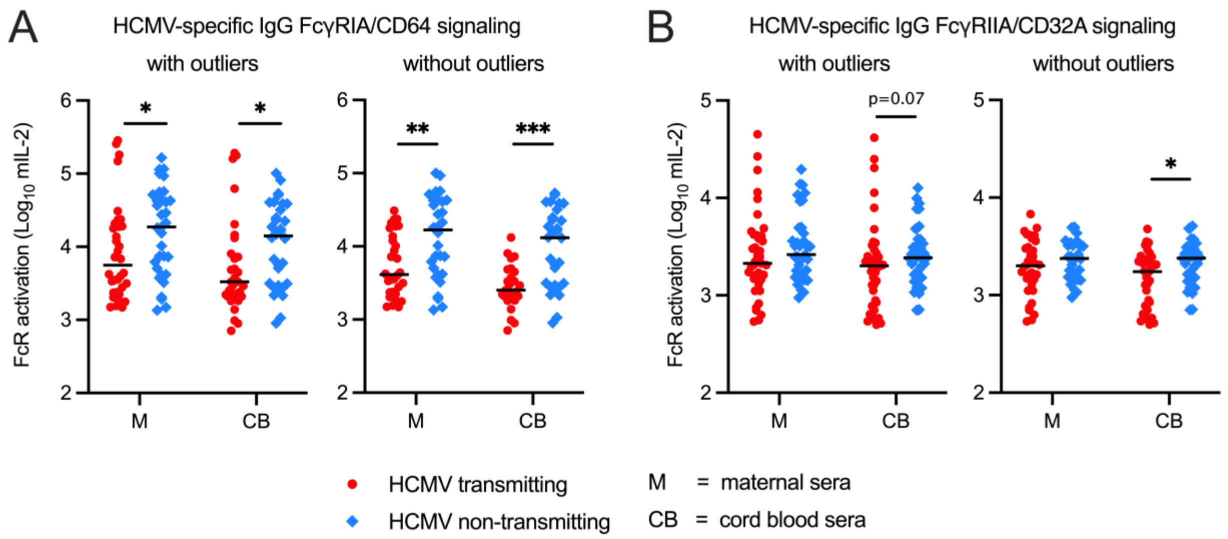


**Supplementary Figure 7. Sensitivity analysis identifies outliers in HCMV-specific IgG activation of FcγRIA/CD64 and FcγRIIA/CD32A in HCMV transmitting and non-transmitting dyads.** Measurements of virus:sera immune complex activation of host FcγRs was analysed for outliers using the ROUT method (Q=1%). (A) HCMV-specific FcγR activation in HCMV transmitting (red circles) versus non-transmitting (blue squares) dyads including outliers. (B) HCMV-specific FcγR activation in transmitting (red circles) versus non-transmitting (blue squares) dyads excluding outliers. P values for Mann-Whitney *U* test. * P < 0.05, ** P < 0.01, *** P < 0.001.

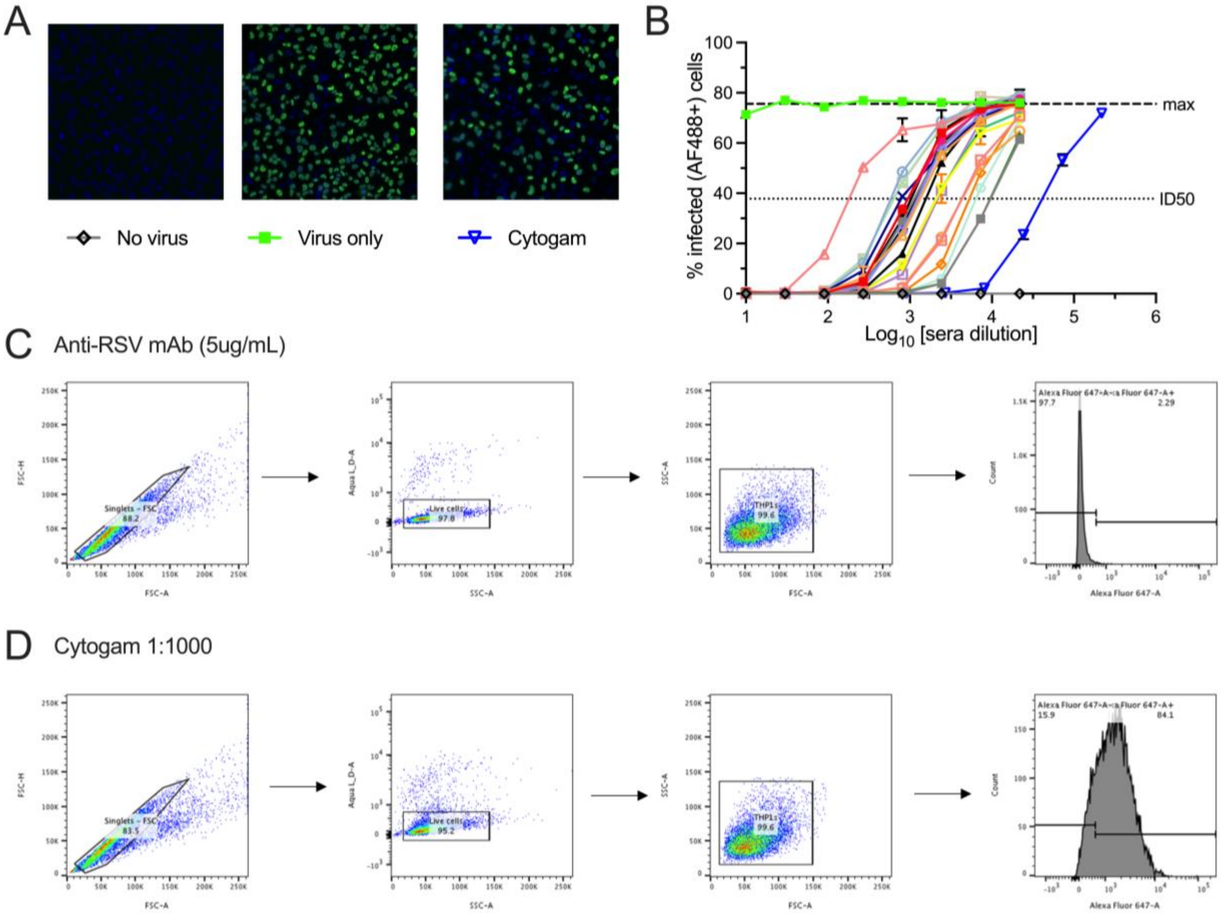


**Supplementary Figure 8. HCMV neutralization and whole virion antibody-dependent cellular phagocytosis (ADCP) assay methods.** Neutralization was measured by HCMV IE1 staining with AF488 and antibody titers were calculated as the inhibitory dilution 50 (ID50), equivalent to the sera dilution that inhibited 50% of the max infection in virus only wells. Infected cell percentage was calculated as proportion of AF488 positive cells out of total DAPI+ cells per well. (A) IE1 staining (green) and DAPI staining (blue) of epithelial (ARPE) cells infected with HCMV AD169r strain showing representative no virus, virus only, and virus plus HCMV-specific hyperimmunoglobulin (Cytogam) treated wells. (B) Plot showing how ID50 values were interpolated from sera serial dilutions. (C-D) Flow cytometry gating strategy for ADCP assay showing gating for (C) Pavalizumab, a non-HCMV-specific monoclonal antibody (mAb) against respiratory syncytial virus (RSV) F protein, (negative control) and (D) Cytogam (positive control).
